# Insights into *KIF11* pathogenesis in Microcephaly-Lymphedema-Chorioretinopathy syndrome: a lymphatic perspective

**DOI:** 10.1101/2023.11.02.23297090

**Authors:** Kazim Ogmen, Sara Dobbins, Ines Martinez-Corral, Rose Yinghan Behncke, Ryan C.S. Brown, Sascha Ulferts, Nils Rouven Hansmeier, Ege Sackey, Ahlam Alqahtani, Christina Karapouliou, Dionysios Grigoriadis, Michael Oberlin, Denise Williams, Arzu Ekici, Kadri Karaer, Steve Jeffery, Peter Mortimer, Kristiana Gordon, Benjamin M. Hogan, Taija Mäkinen, René Hägerling, Sahar Mansour, Silvia Martin-Almedina, Pia Ostergaard

**Author notes:** Current affiliation: University of Lille, Inserm, CHU Lille, Lille Neuroscience and Cognition, Laboratory of Development and Plasticity of the Neuroendocrine Brain, UMR-S, Labex DistAlz, Lille, France. Current affiliation: Developmental Biology and Cancer Department, Great Ormond Street Institute of Child Health, University College London, London, UK. Clinical correspondence to Sahar Mansour, South West Thames Centre for Genomics, St George’s University of London, Cranmer Terrace, London SW17 0RE, United Kingdom. Research-related correspondence to Silvia Martin-Almedina, Molecular and Clinical Sciences Institute, St George’s University of London, Cranmer Terrace, London SW17 0RE, UK.

## Abstract

Pathogenic variants in kinesin *KIF11* underlie microcephaly-lymphedema-chorioretinopathy (MLC) syndrome. Although well known for regulating spindle dynamics ensuring successful cell division, the association of *KIF11* (encoding EG5) with development of the lymphatic system, and how *KIF11* pathogenic variants lead to lymphatic dysfunction and lymphedema remain unknown.

Using patient-derived lymphoblastoid cells, we demonstrate that MLC patients carrying pathogenic stop-gain variants in *KIF11* have reduced mRNA and protein levels. Lymphoscintigraphy showed reduced tracer absorption, and intestinal lymphangiectasia was detected in one patient, pointing to impairment of lymphatic function caused by *KIF11* haploinsufficiency.

We reveal that *KIF11* is expressed in early human and mouse development with the lymphatic markers VEGFR3, Podoplanin and PROX1. In zebrafish, scRNA-seq identified *KIF11* specifically expressed in endothelial precursors. In human lymphatic endothelial cells (LECs), EG5 inhibition with Ispinesib, reduces VEGFC-driven AKT phosphorylation, migration and spheroid sprouting. *KIF11* knockdown reduces *PROX1* and *VEGFR3* expression, providing for the first time a link between *KIF11* and drivers of lymphangiogenesis and lymphatic identity.

## Introduction

Microcephaly-lymphedema-chorioretinopathy syndrome (MLC; ORPHA:2526), also known as ‘microcephaly with or without chorioretinopathy, lymphedema, or intellectual disability’ syndrome, is an autosomal dominant condition with about 50% of patients presenting with congenital primary lymphedema^1^. Variants in *KIF11* cause 75% of MLC cases^1–3^. The pathogenic basis of *KIF11*-associated retinal vascular disease has been investigated through the generation of a mouse model for familial exudative vitreoretinopathy (FEVR)^4^. How *KIF11* mutations lead to lymphedema, the specific hallmark of lymphatic dysfunction, is not known.

*KIF11* encodes EG5, a motor protein essential in cell cycle dynamics. EG5 inhibition arrests cells in metaphase inducing monopolar spindles *in vitro*^5,6^. No phenotype was found in heterozygous mice (Eg5^+/−^), but homozygous deletion (Eg5^−/−^) was lethal in the embryo^7,8^.

Recently, several groups found roles for EG5 independent of cell division, e.g. cell migration, intracellular vesicle trafficking or regulation of primary cilia dynamics in several cell models including post-mitotic neurons^9–13^. These studies could explain the pleiotropic and variable phenotype characteristic of MLC syndrome, potentially related to the diverse functions of EG5, which may even be cell type dependent.

Here we investigate the clinical features and disease mechanisms in MLC patients with *KIF11* pathogenic variants, focusing on lymphatic function. They typically presented with congenital onset lower limb lymphedema, mostly in the dorsum of the foot. Clinically their phenotype resembles that of Milroy disease and lymphoscintigraphy scans showed functional aplasia. Significant issues with intestinal lymphangiectasia were noted in one of the individuals, which has been only reported once in MLC patients^14^, but is characteristic of other primary lymphatic anomalies^15,16^.

We hypothesise that there is a critical role for *KIF11* (EG5) in the lymphatics, possibly through interactions with VEGFR3. We show that *KIF11* co-expresses with *VEGFR3* in the initial steps of lymphangiogenesis in early human and mouse embryonic development. In zebrafish *kif11* expression is associated with endothelial progenitors. *In vitro* loss-of-function models show that *KIF11* inactivation decreases proliferation, migration and sprouting after VEGFC treatment of serum-starved LECs. Here we provide first insights into the association of EG5 with lymphatic development and function in health and disease. More research into the specific roles of EG5 at defined lymphatic developmental stages is required to characterise disease mechanisms and delineate target treatments for MLC patients.

## Materials and methods

### Patient recruitment and ethical approval

Six index cases and affected family members with variants in *KIF11* were included in the study. Two were previously described^3^ and four were direct referrals from clinicians. *KIF11* variants were detected by exome sequencing in the respective molecular genetics services. Ethical approval was given by the South West London Research Ethics Committee (REC Ref: 05/Q0803/257 and 14/LO/0753) and by EA4/214/19 (‘Molekulare und Moprhpogenetische Mechanismen der Lymphödementwicklung’) granted to RH, Charité Ethics committee. Written informed consent to take part in the study and to have the results of this research work published was obtained from all participants or their legal representatives.

### DNA and RNA/cDNA Analysis of the Splice Variant

To analyse the c.2922G>A variant reported in three of the index cases peripheral blood was obtained from individual F5-II.1. DNA was extracted using a standard chloroform ethanol procedure. Sequencing of DNA was performed using *KIF11* (NM_004523.4) forward primer 5’-caggtagcaagactgatcctca-3’ and reverse primer 5’-cggggtaagattgaggggta-3’ covering exon 20. RNA was collected using PAXgene Blood RNA tubes (PreAnalytiX, Hombrechtikon, Switzerland) and total RNA extracted using the PAXgene blood RNA Kit (PreAnalytix). DNAse treatment was performed to eliminate genomic DNA and cDNA synthesized from the RNA with a SuperScript II Reverse Transcriptase using about 400 ng RNA and 50 ng random primers (Invitrogen, Carlsbad, CA, USA). Sequencing of cDNA was performed using primers which span exons 19 to 22 of *KIF11* (NM_004523.4): Forward 5’- GGCAGCTCATGAGAAACAGC-3’ and Reverse 5’-GCGAGCCCAGATCAACCTTT-3’. All PCR products were sequenced using BigDye Terminator v3.1 chemistry (Life Technologies, Carlsbad, CA, USA) and an ABI3130xla Genetic Analyzer (Life Technologies). Sequencing traces were visually inspected in FinchTV v1.4 (Geospiza). The protein prediction tool Expasy (www.expasy.org) was used to confirm the effect of the splice-site variant on the protein sequence.

### Generation of lymphoblastoid cell lines from MLC patients and analysis of KIF11 (EG5) expression levels in blood and lymphoblastoid cell lines

Four patients from families F1 and F2 with MLC presenting with congenital, bilateral pedal lymphedema and two controls were selected for the generation of lymphoblastoid cells to analyse *KIF11* expression. Epstein-Barr virus (EBv) transformation was used for the generation of lymphoblastoid cell lines from the patients’ peripheral blood lymphocytes at the Culture Collections, Public Health England, Porton Down. For the analysis of *KIF11* expression levels, qPCR was performed in total RNA samples extracted from blood and saliva. Extraction of RNA from blood samples was performed following the instructions of the Paxgene Blood RNA kit (PreAnalytiX 762154). Samples were eluted in 80µl of elution buffer provided by the kit and stored at −80°C until analysis. Extraction of RNA from saliva samples was done following the Oragene (OGR-500) collection protocol followed up from the Qiagen microkit RNA kit. RNA quality and concentration were assessed using Nanodrop. All RNA obtained was reverse transcribed using oligo dT (SuperScript III First-Strand Synthesis System 18080-051, Invitrogen). Gene expression levels were analysed using Platinum SYBR Green qPCR super mix-UDG with ROX reagent (QIAGEN) and a 7900 HT Fast Real-Time PCR System thermocycler (Applied Biosystems) following the manufacturer’s instructions. Relative gene expression levels were normalized to GAPDH. Specific primers designed for each of the mutations (underlined bases showing nucleotide changes) plus primers for wildtype and mutant alleles are: L347Efs*8: CTCTCAATCTTGAGGAAACT<u>C</u> / CTCTCAATCTTGAGGAAACT<u>G</u> / CTGATTCACTTCAGGCTTATTC; R387*: CGGGCTGCAGCAAGATCTC<u>G</u> / CGGGCTGCAGCAAGATCTC<u>A</u> / CTGAAGTGAATCAGAAACTC.

Validation of the qPCR was performed using cDNA in which the R387* mutation was introduced by site-directed mutagenesis with the QuickChange XL site-directed mutagenesis kit (Stratagene 200517). Sequences were verified by Sanger sequencing analysis.

Lymphoblastoid cells lines were cultured in suspension in RPMI 1640 supplemented with 2mM Glutamine and 20% Foetal Bovine Serum (FBS). Two independent EBv B cell lines were used as control (a gift from Dr. Nancy Hogg, Cancer Research UK London Research Institute, now part of the Francis Crick Institute). Eg5 monoclonal antibodies against the N-terminal (CC10014, Cell Applications or NB500-181, Novus Biologicals) were used in the western blots and β-Actin was used as an internal control.

### Isolation of intestinal biopsy from an MLC patient and analysis of lymphatic endothelial markers

An MLC patient presenting with failure to thrive, dysphagia and intermittent diarrhea and vomiting was subjected to endoscopy and intestinal biopsies taken for analysis by the Pathology Department at his local hospital. Subsequent immunofluorescence staining for epithelial and endothelial markers on 5µm tissue sections was performed according to the standard immunofluorescence histology protocol. Briefly, fixed tissue sections were washed and blocked (10% chicken serum, 0.3% Triton X-100 in PBS). Following blocking, tissue sections were incubated for 1 hour with primary antibodies (diluted in 1% BSA, 1% chicken serum, 0.3% Triton X-100 in PBS), washed thrice in PBS-T (0.1% Tween20 in PBS) and finally incubated in Alexa dye–conjugated secondary antibodies (Life Technologies). After sample mounting in Mowiol (Calbiochem, 475904), samples were imaged using a Zeiss LSM 980 confocal microscope (25x oil, NA = 0.8).

### RNAscope in situ hybridization in human fetal tissue

Human embryonic tissue was obtained from the MRC/Wellcome-Trust funded Human Developmental Biology Resource (HBDR, http://www.hdbr.org), with appropriate maternal written consent and approval from the Newcastle and North Tyneside NHS Health Authority Joint Ethics Committee. HDBR is regulated by the UK Human Tissue Authority (HTA; www.hta.gov.uk) and operates in accordance with the relevant HTA Codes of Practice. The human tissue was collected at developmental stages ranging from CS12 to CS22. Tissues were primarily fixed in 10% PFA overnight at room temperature, and then secondarily fixed in Methacarn before they were processed using automated Excelsior AS (Thermo Scientific, A823-1005) to generate FFPE blocks. Sections were cut from the blocks at 8 µm using a manual rotary microtome (Leica microsystems RM2235, UK). Simultaneous RNA *in situ* hybridization was carried out using RNAscope Multiplex Fluorescent Reagent Kit v2 (Cat. No. 323100 and 323270, ACD) following the manufacture’s protocol for FFPE tissue mounted on slides, with pre-treatment steps of Hydrogen Peroxide and Protease reagents (Cat. No. 322381, ACD) and target retrieval reagents for 20mins at 95°C (Cat. No. 322000, ACD). The following RNAscope probes were designed and purchased from ACD: Hs-PDPN (Cat No. 539751), Hs-PROX1 (Cat No. 530241), VEGFR3 (Hs-FLT4) (Cat No. 552441-C2), and Hs-KIF11 (Cat No. 562641-C3). The probes were fluorescently labelled using OPAL 520 (Cat. No. FP1487001KT, PerkinElmer), OPAL 570 (Cat. No. FP1488001KT, PerkinElmer) and OPAL 650 dyes (Cat. No. FP1496001KT, PerkinElmer) for direct visualization under automated laser-scanning microscopy (Zeiss CellDiscoverer7, Carl Zeiss, Germany).

### Mouse embryo collection, histology, immunofluorescence and RNAscope in situ hybridization

See Supplementary Materials and Methods for details.

### Zebrafish husbandry, genome editing, imaging and quantification

Zebrafish work was conducted in compliance with animal ethics committees at the Peter MacCallum Cancer Centre and The University of Melbourne. The following transgenic lines were used: Tg(*fli1a:nEGFp*)*^y^*^7^ ^17^ and Tg(*-5.2lyve1b:dsred2*)*^nz101^* ^18^. A guide RNA (gRNA) target was designed using CHOPCHOP^19^ to target a highly conserved sequence within Exon 9 of the *kif11* gene and ordered from Integrated DNA Technologies (IDT). gRNA preparation was completed as previously described^20^. ∼250ng/ul of the gRNA were co-injected with 0.5ug of Cas9 protein to AB. F0 adults were outcrossed and germ line transmission of the mutation were selected for using genotyping primers. Transmitting F0 fish were outcrossed and F1 adults were selected to stabilise the 8bp mutation based on homozygous phenotype genotyping. gRNA oligo: 5’- taatacgactcactata**GGTGAGAACATCGGACGATC**GTTTTAGAGCTAGAAATAGC-3’.

Genotyping primers: 5’-GCTTTCGAGTCAGTGGGGTT-3’, 5’- CCTGCAGTATGCGGGTTAGT-3’. Embryos were mounted in 0.5% low meting agarose (Merck, Darmstadt, Germany; A9414-100G) as previously described^21^. Imaging was completed at the Centre for Advanced Histology and Microscopy (The Sir Peter MacCallum Cancer Centre). Images in **Fig 5f** were taken on an Olympus BX53 microscope (Differential inference contrast (DIC) imaging, 10x Objective). Images in **Fig 5g-i** were taken on a Nikon SoRa Spinning Disk Confocal, the 10x objective was used for brightfield images, and 20x objective used for fluorescent imaging in **g & i**. Fluorescent images were stitched together to visualise the whole embryo. Images in **Fig 5i** were taken using an Olympus Yokogawa CSU-W1 spinning disc confocal microscope (brightfield, 10x objective & fluorescent imaging, 20x objective). Endothelial cells (*fli1a:nEGFP*) were manually quantified with FIJI, Image J (National Institutes of Health)^22^. Statistics were completed for these images as previously described^23^.

### Zebrafish scRNA-seq data processing and analysis

The previously published scRNA-seq data set^24^ was processed as described in the original publication^23^. Downstream analysis and visualisation were performed in R (v4.2.0) using built in functions in Seurat (v3.0). Pair-wise gene spearman correlations were calculated with log normalised gene counts using the cor.test function from the stats package.

### Lymphatic endothelial cell culture, siRNA transfection and Ispinesib treatment

Human dermal LECs (C-12217, PromoCell) were cultured and maintained in endothelial cell growth medium MV2 (C-22022, PromoCell) with FCS-based supplement mix (C-39226) and recombinant human VEGFC 50ng/ml (2179-VC-025, R&D Systems). Transfection of cells with siRNA *KIF11* (NM_004523, SI02653693, target sequence 5’- ACGGAGGAGATAGAACGTTTA-3’) or siRNA ‘All starts’ negative control (1027281, Qiagen) was performed with Lipofectamine 2000 (Life Technologies) following the manufacturer’s recommendations. Ispinesib (Tocris), a specific EG5 antagonist, was used at various concentrations and exposure times depending on the nature of the experiment. DMSO was used as vehicle.

### Lymphatic endothelial cell functional assays

LECs were used in various functional assays such as transwell migration, wound healing, single cell migration tracking and spheroid sprouting assays. See Supplementary Materials and Methods for details.

### Tyrosine Kinase (TK) array and Bioinformatic Analysis

The Human Phospho-Kinase Array (#ARY003B, R&D systems) was hybridised following the manufacturer’s recommendations. Briefly, membranes containing captured antibodies against 43 kinases in duplicate, were incubated with either siRNA control or siRNA *KIF11* treated LECs lysates collected 24 hours post-transfection. Chemiluminescence was used for the detection of antigen-antibody complexes, generating images for different exposure times. Signals were quantified using Image J^25^ and results plotted with GraphPad Prism version 9.5.1 for Windows.

For the two duplicates the average Fold Change (FC) and the average Coefficient of Variation (CoV) was calculated for the normalised control and treatment levels for each kinase. Kinases were selected as dysregulated where FC>1.1 or FC<0.9, and where the average CoV was smaller compared to the observed variation between treatment and control, (plus FC effect must be twice as large as the average CoV for that kinase) at either the 1min or 10min chemiluminescence exposure timepoints. Selected kinases were excluded which were not consistent in the direction of change across all time points analysed. Interactions between the dysregulated kinases were identified using STRING (v11.5)^26^ based on experimentally verified interactions. Cytoscape (v3.8.2)^27^ was used to visualise the protein networks.

### Statistical analysis

Prims 9.0 software (GraphPad) was used for all statistical assessments apart from the single cell migration tracking done with R. Statistical significance between two groups was assessed by unpaired parametric two-tailed t-test. Statistically analysed data are presented as mean ± SEM or mean ± SD as specified for each analysis (described in the Figure legends). Differences were considered significant when P < 0.05.

## Results

### MLC patients show lymphatic functional defects

Lymphedema in MLC patients is mostly bilateral, affecting lower limbs^1,3^. We evaluated nine patients with an MLC phenotype, five individuals from four families carrying two different unreported *KIF11* variants: c.2680C>T; p.(Q894*) and c.2922G>A; p.(P974=), and four individuals from two previously reported families^3^ carrying pathogenic variants in *KIF11,* c.1159C>T; p.(R387*) and c.1039_1040delCT; p.(L347Efs*8). Clinical summary is in **Table S1**. Pedigrees are shown in **Fig. S1**.

Lymphatic function of the lower limbs was assessed in three individuals using lymphoscintigraphy. Proband F1-II.1, who presented with congenital bilateral lymphedema, showed absence of radioactive isotope uptake from the web spaces between the toes (i.e. functional aplasia) as described previously^3^ (**Fig. 1a**). Two siblings, F3-II.1 and F3-II.2, presenting with microcephaly and congenital bilateral lower limb lymphedema also demonstrated abnormalities (**Fig. 1b-c**). One leg is more affected than the other in both siblings, but with evidence of functional aplasia in one leg the findings are in keeping with an underlying bilateral primary lymphedema compared to the control (**Fig. 1d**).

**Fig. 1:**
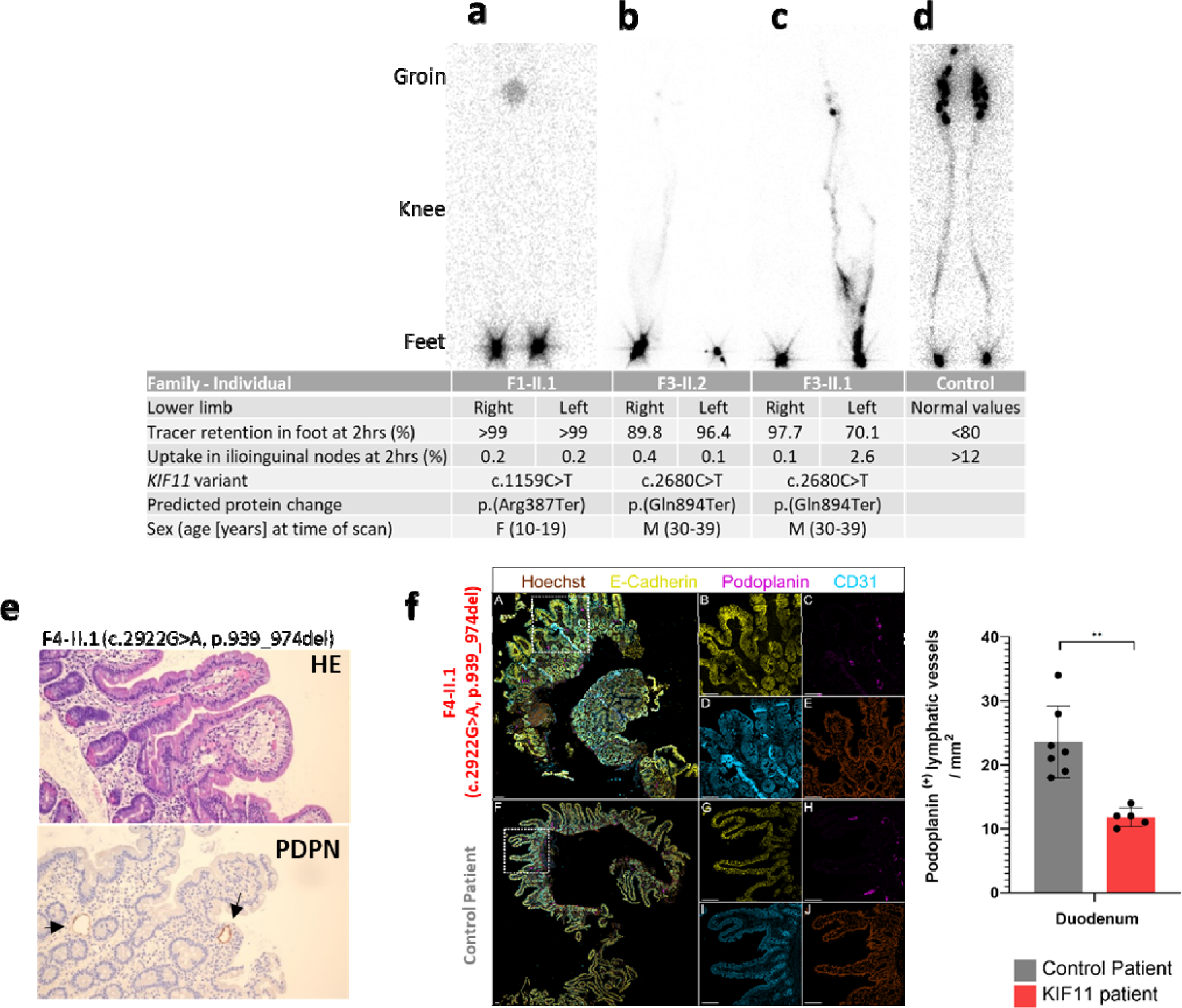
MLC patients show lymphatic abnormalities. **(a-d)** Lower limb lymphoscintigraphy 2 hours after injection is used to image the lymphatic system (anterior view shown here). Quantification figures, and genetic variants and predicted protein changes are shown in table below. **(a)** Bilateral functional aplasia evidenced by no lymphatic drainage in either leg, and quantification of the tracer signal 2 hours post-injection revealed that almost 100% of activity was retained in both feet with a reduced intake in the groin of only 0.2%. **(b)** Significantly reduced function with rerouting on the right and functional aplasia on the left. **(c)** Functional aplasia on the right and abnormal tortuous tracts with patchy superficial re-routing on the left. **(d)** Unaffected subject with symmetrical transport of radionuclide tracer from injection sites in the feet up to the inguinal lymph nodes via main lymphatic vessels. Values for an individual with a normal lymphatic system are given in the table. **(e)** Intestinal biopsy of patient F4-II.1 with *KIF11* variant c.2922G>A; p.939_974del. H&E (HE) staining shows aberrant morphology of the lacteals, Podoplanin (PDPN) detected slightly enlarged lymphatic vessels (arrows). **(f)** Human duodenum sections derived from a *KIF11* patient (F4.-II.1 c.2922G>A; p.939_974del.) and an unaffected patient were subjected to immunofluorescence staining. Selected magnifications (framed areas) depict staining for Hoechst (brown; E, J), E-cadherin (yellow; B, G), Podoplanin (magenta; C, H), CD31 (cyan; D, I). Scale bars represent 100 µm. Immunofluorescence staining revealed significantly reduced lymphatic vessel density compared to healthy control samples based on Podoplanin positive area per mm^2^ tissue area. Two tailed Student t-test p<0.01.

Proband F4-II.1 presented with dysphagia, intermittent diarrhoea, and failure to thrive; a clinical observation not typical of MLC. Podoplanin staining of intestinal biopsies showed dilated lymph vessels (lacteals) in the duodenum (**Fig. 1e**), and together with the significantly increased Alpha 1 antitrypsin (A1AT)-values in the stool, is consistent with protein-losing enteropathy (PLE) caused by intestinal lymphangiectasia. Immunofluorescence staining revealed reduced lymphatic vessel density compared to healthy control samples based on Podoplanin positive area per tissue area (**Fig. 1f**).

### MLC patients show reduced expression of KIF11 (EG5) pointing to haploinsufficiency as the disease mechanism

To ascertain the disease mechanism in MLC, we analysed the expression of *KIF11* in lymphoblastoid cell lines, blood and saliva from four patients carrying pathogenic variants (leading to a premature termination) in the motor domain (L347Efs*8) and the coiled-coil domain 1 (R387*) (**Fig. 2a**). These regions are involved in protein oligomerisation^28^ and vesicle tethering^29^ respectively, and thus seem likely that both variants will negatively impact on EG5 function. qRT-PCR analysis of blood (**Fig. 2b**) and saliva (data not shown) using allele-specific primers for the wild-type (wt) allele and each of the mutant alleles (mut) showed strong reduction, but not a complete absence of the mutant mRNA in all samples. This fits with nonsense mediated RNA decay, and the then expected level of wt *KIF11* mRNA of 50% in patients’ samples compared to the controls. Western blot analysis of protein lysates from lymphoblastoid cell lines also showed around 50% levels of EG5 protein compared to the controls (**Fig. 2c**). Interestingly, a truncated protein of expected size (43KDa) was detected in F1-II.1 (**Fig. 2d**).

**Fig. 2:**
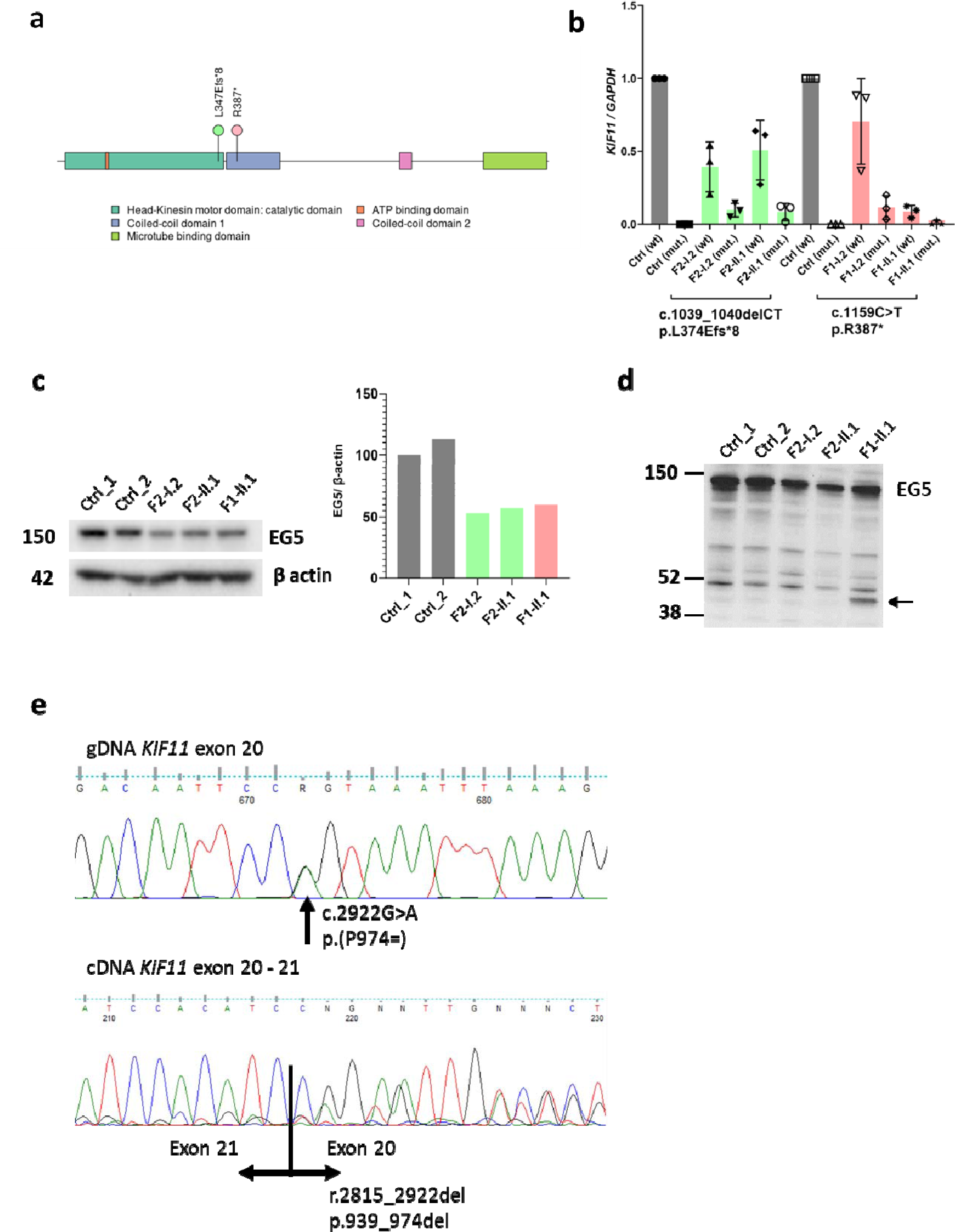
Reduced expression of *KIF11* (EG5) and splicing abnormalities suggest haploinsufficiency as the disease mechanism in MLC. **(a)** EG5 (*KIF11*) protein domain structure indicating the position of the MLC mutations p.L347Efs*8 and p.R387* for which patient-derived lymphoblastoid cells and blood samples were available. **(b)** qRT-PCR analysis of blood using allele-specific primers (wt for wild type allele and mut for allele containing the mutation) showing strong reduction, but not a complete absence of the mutant mRNA in all analysed samples (F2-I.2, F2-II.1, F1-I.2 and F1-II.1), suggesting nonsense mediated RNA decay. wt *KIF11* mRNA levels were decreased by up to 50% in patients compared to the controls. Data represent average relative expression (n=3 experiments) ± SD. **(c)** Western blot analysis of protein lysates from lymphoblastoid cell lines also indicate approximately 50% reduction in the levels of wt KIF11 protein (119kDa) in the patient samples (F2-I.2, F2-II.1, and F1-II.1) using C-terminal binding EG5 antibody (NB500-181, Novus Biologicals). Experiment shows an average of 2 technical repeats of identical biological materials. Beta-actin was used as a loading control. **(d)** Western blot analysis with an N-terminal binding anti-EG5 antibody (CC10014, Cell Applications, USA) showed the presence of a truncated protein of the expected size in patient F1-I.2 (arrow). The position of molecular mass markers (in kDa) is indicated on the left of the gel. **(e)** Top panel: Sanger sequencing of gDNA from proband F5-II.1. PCR product shows a compound peak in exon 20, which is the synonymous *KIF11* c.2922G>A; p.(P974=) variant. Lower panel: Sanger sequencing of cDNA from F5-II.1. Black line indicates the boundary between the last base of exon 20 and first base of exon 21. Notice the heteroduplex in exon 20 indicating exon skipping. Analysis of the full trace identified a loss of the last 108 bases in exon 20 (r.2815_2922del) which is predicted to lead to a shorter EG5 protein similar to that of the synonymous variant c.2922G>T as shown by others^50^.

### A synonymous variant in KIF11 causes cryptic splicing which leads to exon skipping

Three MLC patients in this study carry the same synonymous variant c.2922G>A; p.(P974=) (**Table S1; Fig. 2e**) and *in silico* prediction tools suggest the variant is pathogenic (**Table S2**). RNA was isolated from patient F5-II.1, converted into cDNA and Sanger sequenced. The synonymous variant causes cryptic splicing which leads to the removal of a 108bp fragment of exon 20 (r.2815_2922del) possibly leading to a shortened EG5 protein (p.V939_P974del) (**Fig. 2e**).

### KIF11 co-expresses with lymphatic VEGFR3 positive structures during human embryonic development

*In silico* analyses show kinesin transcripts, including *KIF11*, are enriched in the endothelial expressed sequence tag (EST) pool^30^, and human foreskin sections showed positive co-staining of EG5 with CD31 and Podoplanin, confirming the expression of EG5 in adult lymphatics. As MLC is characterised by early onset lymphedema there is a need to understand *KIF11* expression patterns during lymphatic embryonic development. Human embryonic tissues at different developmental stages ranging from human Carnegie stage 12 (CS12) to CS22 were analysed for *KIF11* mRNA expression along with different lymphatic specific markers (*VEGFR3* and *PROX1* or *PDPN*) using RNA-scope multiplexing *in-situ* hybridization. Although co-expression of *KIF11*, *PROX1* and *VEGFR3* was found in the hepatic primordia (HP), little or no co-expression of *KIF11* with other lymphatic markers was seen around the developing cardinal vein (CV) at CS12 (**Fig. 3a**). Later at CS15, low expression levels of *KIF11,* with co-expression to *PROX1* and *VEGFR3,* was detected around the CV. At CS18, increased expression of *KIF11* with *VEGFR3* and *PDPN* positive cells was found around the CV. These VEGFR3 positive cells would migrate from the cardinal vein to form the primordial thoracic duct (pTD), formerly known as the lymphatic sacs^31^. Later in development, at CS22, the association between *KIF11* and lymphatic markers was maintained as the lymphatic vessel network expanded. This indicates that *KIF11* is associated with the development of lymphatic vessels at the later stages analysed.

**Fig. 3:**
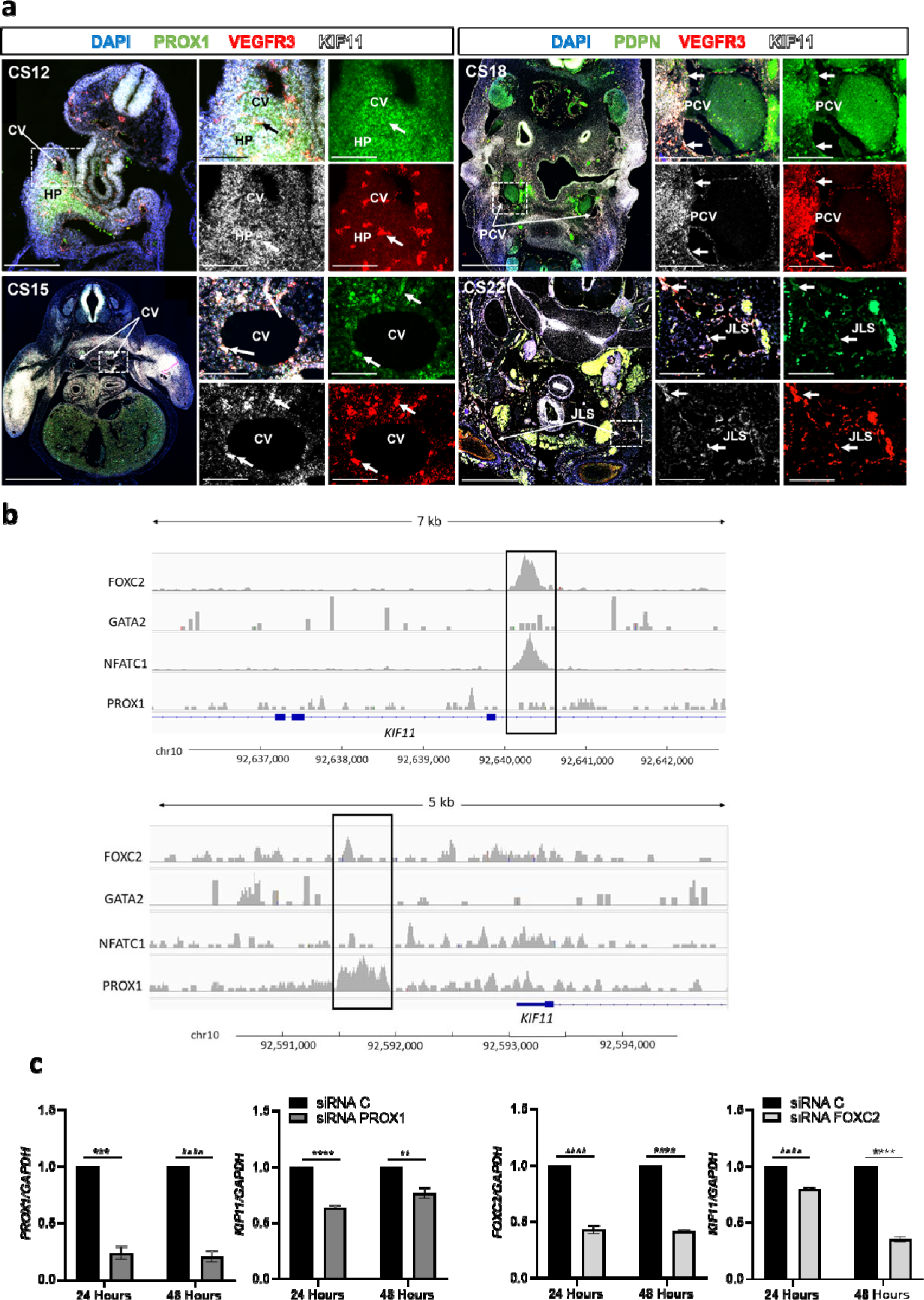
*KIF11* expression during human embryonic development and transcriptional control in adult lymphatic endothelial cells. **(a)** Human embryonic slides corresponding with different embryonic developmental Carnegie stages (CS12, CS15, CS18 and CS22) were hybridised with fluorescence probes for *KIF11* (white), *PROX1* or *PDPN* (green) and *VEGFR3* (red). Magnified regions marked with a square box in overview images contain the pre-cardinal veins (PCV), cardinal veins (CV) and jugular lymphatic sacs (JLS), and arrows point to regions of co-expression between *KIF11* and lymphatic endothelial marker expression. HP, hepatic primordia. Scale bar 200 µm. **(b)** ChIP-seq analysis in cultured human dermal LECs identifies an intronic region of *KIF11* bound by FOXC2 and NFATC1 (top panel) and PROX1 binding 1.3kb upstream of *KIF11* promoter (bottom panel). **(c)** LECs were treated with siRNA PROX1 or siRNA FOXC2 and KIF11 expression was evaluated by qPCR. Bars represent mean relative expression ± SEM. from n=3 experiments. **P<0.01, ***P<0.001, ****P<0.0001. Two-tailed unpaired Student’s t-test.

### KIF11 expression might be regulated by lymphatic transcription factors in human lymphatic endothelial cells

To understand the relationship between *KIF11* expression and key transcriptional regulators of lymphatic development, we analysed publicly available ChIP-seq data of primary human dermal LECs^32^ to identify regions on *KIF11* bound by GATA2, FOXC2, NFATC1 and PROX1. We identified binding of FOXC2 and NFATC1 in intron 17 (chromosomal position chr10:92,640,060-92,640,600) and binding of PROX1 approximately 1.3kb upstream of the *KIF11* promoter (chr10:92,591,500-92,592,000) (**Fig. 3b**). Both peaks overlap with ENCODE enhancer-like signals predicted from DNase hypersensitivity, histone modification or CT-CF binding, with data combined across cell types^33^. To experimentally confirm that PROX1 and FOXC2 may regulate *KIF11* expression, LECs were treated with either siRNA *PROX1* or siRNA *FOXC2* and *KIF11* expression evaluated by RT-qPCR, showing downregulation (**Fig. 3c**).

### KIF11 co-expresses with lymphatic VEGFR3 positive structures during mouse embryonic development

In mice, lymphatic vessels develop at around developmental day E10.0. A subpopulation of Prox1-positive cells, so called initial LECs (iLECs), leave the cardinal vein (CV) and form the primordial thoracic duct (pTD)^31^. A second lumenized lymphatic vessel, the peripheral longitudinal lymphatic vessel (PLLV) is formed simultaneously, which represents a potential source or vascular connection site for dermal lymphatic vessels^31^. We therefore investigated the expression of *KIF11* and *VEGFR3* in the CV, iLECs, the pTD and dermal LECs/PLLVs.

First, we located the relevant lymphatic structures by H&E staining (**Fig. 4a, A, F, K, P**) and immunofluorescence (**Fig. 4a, B, G, L, Q**) on mouse embryonic slides from E10.5 to E13.5. Using RNAscope, we identified a very weak *VEGFR3* mRNA signal in the CV (**Fig. 4a E1 and E4**), but robust *VEGFR3* mRNA signal in iLECs (**Fig. 4a D1 and D4**) at E10.5. From E11.5-E13.5 when the formation of the pTD/lymph sac begins, there is an increased intensity of expression of *VEGFR3* in the cells lining the structure (**Fig. 4a J1 and J4, O1 and O4, T1 and T4,** and **Fig. 4b**). This increase of signal in developing and expanding lymphatic vessels is consistent with the finding in humans at later developmental stages (e.g. CS22, **Fig. 3a**).

**Fig. 4:**
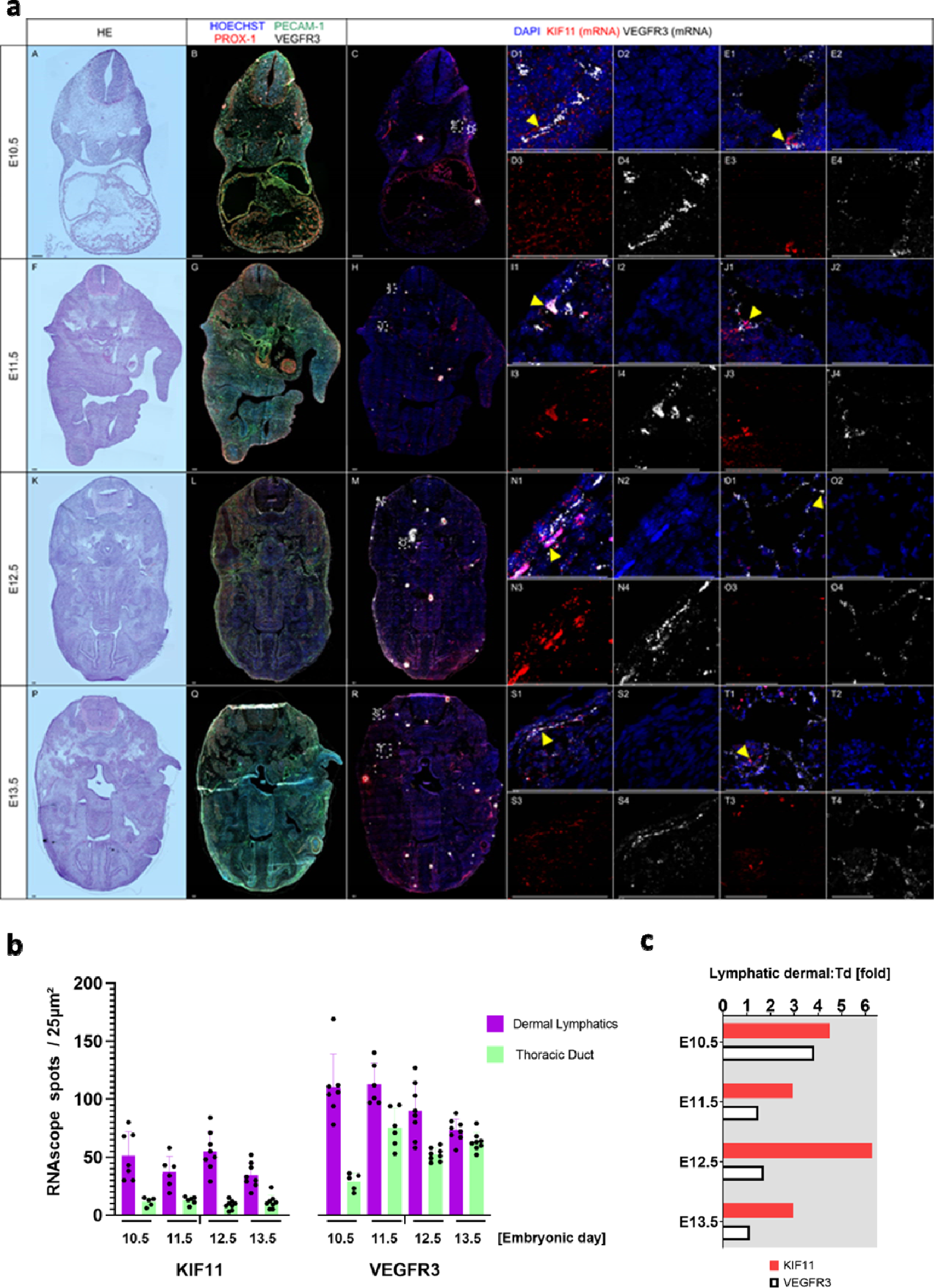
*KIF11* co-expression with *VEGFR3* positive lymphatic vessels during mouse embryonic development. **(a)** Mouse embryonic sections corresponding with different embryonic developmental stages (E10.5 – 13.5) were subjected to H&E staining (A, F, K, P), immunofluorescence staining (B, G, L, Q) or RNA hybridization (C, H, M, R), respectively. Selected magnifications (framed areas) highlight embryonic regions with dermal lymphatics (D, I, N, S) and the thoracic duct (E, J, O, T), which were imaged for DAPI staining (blue; 2) as well as *KIF11* (red; 3) and *VEGFR3* (white; 4) mRNA expression; a merged image is shown in D1, E1, I1, J1, N1, O1, S1, T1. Arrowheads indicate regions with col-expression of *KIF11* and *VEGFR3* mRNA expression. Scale bars represent 100 µm. **(b)** Quantification of *KIF11* and *VEGFR3* mRNA signals from dermal (purple) and thoracic duct (light green) of different stages of mouse embryo development. RNAscope signals were measured from 6-8 different areas of 25um^2^ **(c)** Bar chart represents KIF11 (red) or (white) VEGFR3 (white) mRNA signal ratio in dermal versus thoracic duct (Td) during different stages of mouse embryo development.

*KIF11* mRNA signal was detected to co-express with cells expressing *VEGFR3* from E11.5 to E13.5 in the pTD (**Fig. 4a J1 and J3, O1 and O3, T1 and T3**) at relatively constant levels albeit of a lower signal intensity (**Fig. 4b**). Likewise, dermal cells with a *VEGFR3*-positve mRNA signal express *KIF11* (**Fig. 4a I1 and I3, N1 and N3, S1 and S3**). The *KIF11* mRNA signal appears to be 3- to 5-fold higher in dermal LECs compared to that of the pTD (**Fig. 4c**).

### kif11 expression is associated with highly proliferative lymphatic and blood endothelial precursors during zebrafish embryonic development

To gain further insights into the cellular expression pattern of *KIF11* during development, we analysed recently published scRNA-seq data of wildtype zebrafish cell populations at 3- and 4-days post fertilization (dpf)^24^ (**Fig. 5a**). *kif11* expression was evaluated across all reported cell types (**Fig. 5b**) and compared with the expression of markers for proliferation (**Fig. 5c**). *kif11* expression was positively correlated with expression of the proliferative marker *mki67* in endothelial populations **(Fig. 5d)**.

**Fig. 5:**
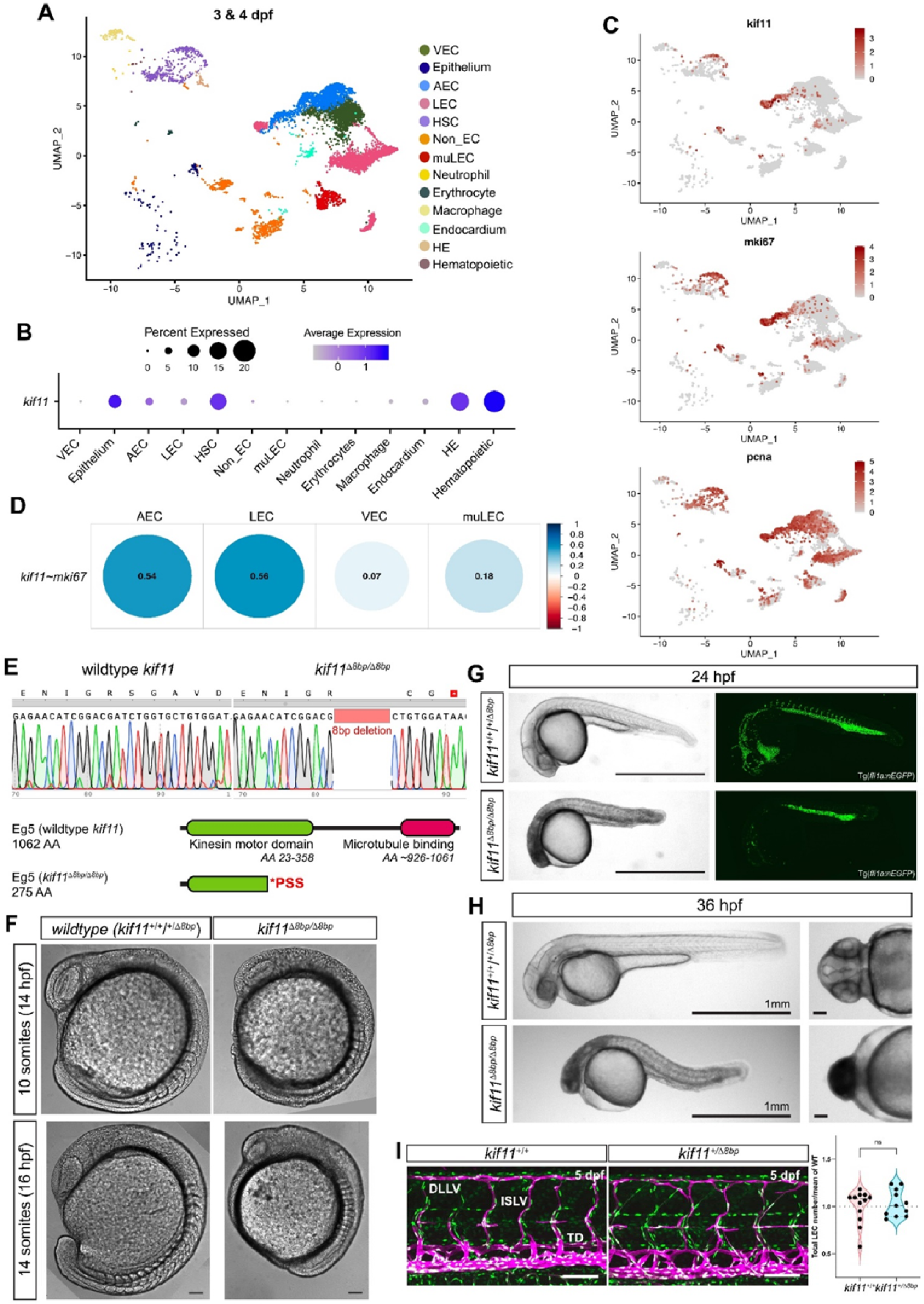
*kif11* expression and function during zebrafish development. **(a)** UMAP visualisation of scRNA-seq data of endothelial cell and related cell populations within wildtype zebrafish at 3 and 4 days post fertilization (dpf). **(b)** Dotplot of *kif11* expression across all reported cell types. Size of circles indicates percentage of cells which express *kif11* and the colour range is shaded from high to low expression. **(c)** UMAP visualisation of *kif11* and proliferation markers *mki67* and *pcna* expression, revealing expression of *kif11* in proliferating cells. **(d)** Dotplot showing Spearman correlation of gene expression between *kiff11* and *mki67* within endothelial cell populations. Size of circle indicates significance value (all correlations p-value < 0.05) and blue to red shading indicates positive or negative correlations, respectively. Value inside dots gives level of correlation. Data from **a**-**d** sourced from Grimm and colleagues ^24^. **(e)** Sequencing reads in wildtype and *kif11*^(8bpdel/8bpdel)^ zebrafish generated using the CRISPR/Cas9 approach (upper panel). The 8 bp deletion induces a frameshift and subsequent premature stop codon. The predicted Eg5 protein structure of the wildtype and *kif11*^(8bpdel/8bpdel)^ zebrafish. The premature stop codon (PSS) induces a truncation within the kinesin motor domain of Eg5 (encoded by *kif11*). (Source: https://www.ebi.ac.uk/interpro/protein/UniProt/F1QK82/). **(f)** Differential inference contrast (DIC) images shows abnormal rostral development in homozygous animals at the 10 somite stage (∼14hpf) and somite defects, neural necrosis and delayed tail extension at 14 somite stage (∼16hpf). Scale = 100µm. **(g)** Brightfield images at 24 hours post fertilization (hpf) show severe necrosis and global developmental delay within homozygous animals. Confocal images of endothelial cells show impaired intersegmental vessel sprouting and loss of endothelial cell in the head. **(h)** Brightfield images at 36 hpf demonstrate developmental delay compared to wildtype siblings. Scale =1mm; or =100µm in the dorsal view of heads in right panel. **(i)** Lateral views of wildtype and *kif11* heterozygous trunk vasculature with endothelial nuclei (green) and veins and lymphatics (magenta) labelled. No difference was observed in lymphatic endothelial cell numbers in *kif11* heterozygotes compared with controls, n=12 wildtype siblings and n=11 *kif11*^(+/8bpdel)^.

Next, to model the pathogenic basis of *KIF11*-associated disease, we generated a zebrafish mutant model using CRISPR-Cas9, a *kif11* knockout harbouring an 8bp deletion *kif11*^(8bpdel/8bpdel)^ in the coding sequence (**Fig. 5e**). Similarly to some pathogenic *KIF11* variants observed in MLC, the deletion induces a frameshift and subsequent premature stop codon, predicted to truncate the kinesin motor domain of Eg5 (**Fig. 5e**). Analysis of wildtype and *kif11*^(8bpdel/8bpdel)^ mutants revealed an early developmental phenotype in homozygous animals at ∼14 hpf (hours post fertilization) characterised by the abnormal rostral development of the brain, and at ∼16hpf demonstrated with aberrant somitogenesis, early indications of neural necrosis and delayed tail extension (**Fig. 5f**). 24 hpf homozygous embryos showed neural necrosis, loss of eyes and global developmental delay (**Fig. 5g**) and 36 hpf embryos still displayed necrosis and were developmentally delayed (**Fig. 5h**). Thus, even with the introduction of a deletion mimicking the variants observed in some MLC patients, we do not see a comparable phenotype in zebrafish. Rather we see severe defects in cell survival and development that lead to early embryonic lethality. These defects are epistatic to analysis of *kif11* function in the development of the lymphatic system. Furthermore, *kif11* heterozygous mutants show no obvious change in lymphatic endothelial cell number (**Fig. 5i**).

### KIF11 inhibition impairs VEGFC driven cell migration and sprouting in LECs

VEGFR3 is the essential signalling protein in maintaining lymphatic function. The similarities between the congenital bilateral lower limb lymphedema and functional aplasia on lymphoscintigraphy imaging seen in Milroy disease (caused by *VEGFR3* mutations) and MLC (caused by *KIF11* mutations), and the co-expression of *KIF11* and *VEGFR3* during human and mouse embryonic development, may suggest a common functional pathway.

With *KIF11* haploinsufficiency confirmed as the disease mechanism, we generated two loss-of-function approaches to block EG5 function: siRNA transfection and a specific EG5 antagonist, Ispinesib, widely used to investigate EG5 function in several cell models including HUVEC^30^. After 24h, siRNA *KIF11* transfection in LECs reduced >80% EG5 protein expression (**Fig. S2a**) with the expected decrease in cell proliferation measured as %EdU^+^ LECs (**Fig. S2b**). Spindle pole formation after Ispinesib treatment (0.5nM-50nM) was evaluated by α tubulin staining (**Fig. S2c**). Ispinesib at 5nM-50nM caused an abnormal distribution of the filament fibers, compared to vehicle treated cells, due to the inhibition of the kinesin motor function necessary for the assembly of the microtubules^34^, verifying the inhibitory effect of Ispinesib on mitotic function of EG5 at the analyzed doses. Toxicity was tested in a spheroid-based assay by measuring fluorescence uptake of a dye by viable cells. 50-100nM Ispinesib treated spheroids showed comparable fluorescence levels to DMSO controls and retained some ability to sprout, whereas 200nM treated spheroids showed increased toxicity as well as impaired sprouting ability (**Fig. S2d**).

To investigate the possible role of EG5 in regulating LEC migration, firstly, we performed *in vitro* transwell migration assays to measure the ability of LECs to migrate towards VEGFC. Ispinesib inhibited migration in a dose-dependent manner and a 50% reduction was seen after 50nM Ispinesib treatment (**Fig. 6a**). With an approximately 50% loss of *KIF11* expression in MLC patients (**Fig. 2b-c**), we sought to use 50nM Ispinesib for further experiments as our haploinsufficiency *in vitro* model.

**Fig. 6:**
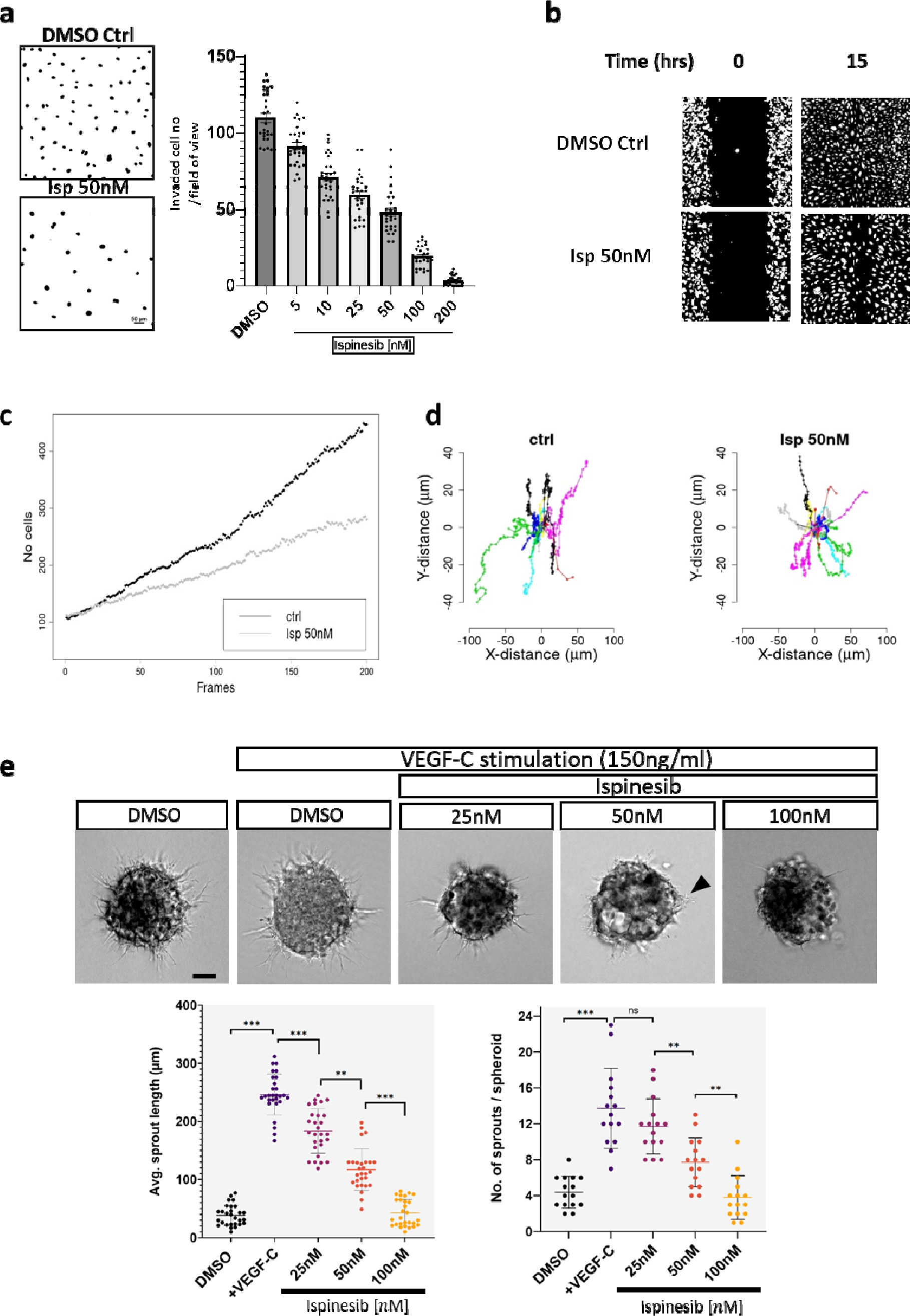
*KIF11* deficiency impairs LEC migration and sprouting. **(a)** 5 x 10^4^ serum starved cells were plated per fibronectin-coated transwell chamber and given 10 hours to migrate in the presence of increasing concentrations of Ispinesib (0 to 200 nM) before fixation and staining of the nuclei with DAPI. Resulting membranes (two examples given on the left) were scored for the movement of nuclei through the transwell membrane, and results represented in a plot (on the right). Error bars represent SD, n = 3 biological replicates (with 2 technical replicates per biological replicate) **(b)** Single cell tracking migration assay follows wound closure ability over 15 hours in DMSO (Ctrl) and 50nM Ispinesib treated cells. **(c)** Number of cells in experimental wound-window area for Ctrl and 50nM Ispinesib treated cells over time (frames). **(d)** Distance and direction of a subset of cells illustrated as star plots of cell tracks for Ctrl and 50nM Ispinesib treated cells. (**b-d**) One representative experiment is shown from n=2. **(e)** 3D spheroid sprouting assay. Sprouting was stimulated in LECs by VEGFC 150 ng/ml either in the presence of DMSO vehicle or several doses of Ispinesib (25,50,100nM). Average sprout length in µm (28 spheroids analysed) and total number of sprouts per spheroid (15 spheroids analysed) were quantified in each condition. One representative image is shown per condition from n=2 biological repeat. ns=not significant, *p<0.05, **p<0.01, ***p<0.001. Two-tailed unpaired Student’s t-test. Scale bar 100 µm.

To further analyze migration parameters in more detail, we performed wound healing experiments tracking single cells moving into the wound. The aim was to study the role of EG5 on cellular migration minimizing interference from cell division, considering that the average full cell cycle length of primary LECs is >20 hours. The wound was fully closed after 15 hours in the control condition, but a delay was observed in 50nM Ispinesib treated cells (**Fig. 6b, Video S1**) as well as a reduction of the number of cells in the experimental wound window over time (**Fig. 6c**). Star plots showed different direction migration patterns between control and Ispinesib treatments **(Fig. 6d**), and velocity analysis indicated a significant reduction of individual cell speed in Ispinesib treated cells with the mean velocity of cells at 2.08 for control vs 1.94 for 50nM Ispinesib treated cells (Wilcoxon-rank-sum test; p-value<2.2×10^−16^).

Lymphatic sprouting is a key process driving lymphangiogenesis in the developing lymphatic system, which requires collective action of both proliferation of stalk and migration of tip cells ^35^. We assessed the role of EG5 in LEC sprouting using an *in vitro* 3D spheroid assay. After 24h, 150ng/mL VEGFC increased the average sprouting length by 60%, and the number of sprouts by 30%, per spheroid compared to the vehicle control. This was reduced in a dose-dependent manner by Ispinesib treatment (**Fig. 6e**). 50nM Ispinesib caused approximately 50% reduction on both the average length and the number of sprouts compared to VEGFC only, and 100nM Ispinesib completely abolished VEGFC-induced sprouting. Overall, the data suggests a role for EG5 in LEC sprouting, probably due to a combination of both the impact on proliferation and migration. To differentially test the effects purely on migration, we attempted to inhibit proliferation prior to Ispinesib treatment, however in our hands, the combination of Ispinesib and aphidicolin (a cell cycle inhibitor) was very damaging for the cells (data not shown).

### KIF11 inhibition reduces VEGFC induced VEGFR3 signalling leading to loss of AKT and MAPK phosphorylation

To understand the impact of *KIF11* haploinsuficiency on lymphatic endothelial cell signalling pathways regulating migration and sprouting we used Proteome Profiles Human Phospho-Kinase Array (R&D) to detect phosphorylation changes in 43 human kinases in siRNA *KIF11* treated LECs compared to siRNA control. Quantification of the signal intensities demonstrated no striking changes in phosphorylation patterns, but we could detect decreased phosphorylation of p38MAPK and AKT1 (at both positions Ser-473 and Thr-308), and an increase in phosporylation of other kinases including JUN and TP53 (**Fig. 7a**), the last associated with spindle defects and stress response^36^. We explored the molecular interactions derived from the human phospho-kinase analysis, plus the already known interactions described in the literature and public databases, by pathway analysis using STRING and Cytoscape. A selection of the kinase networks generated, showing interactions and the direction of the phophorylation changes (blue, decrease of phosporylation; red, increase of phosphorylation) are shown in **Fig. 7b**.

**Fig. 7:**
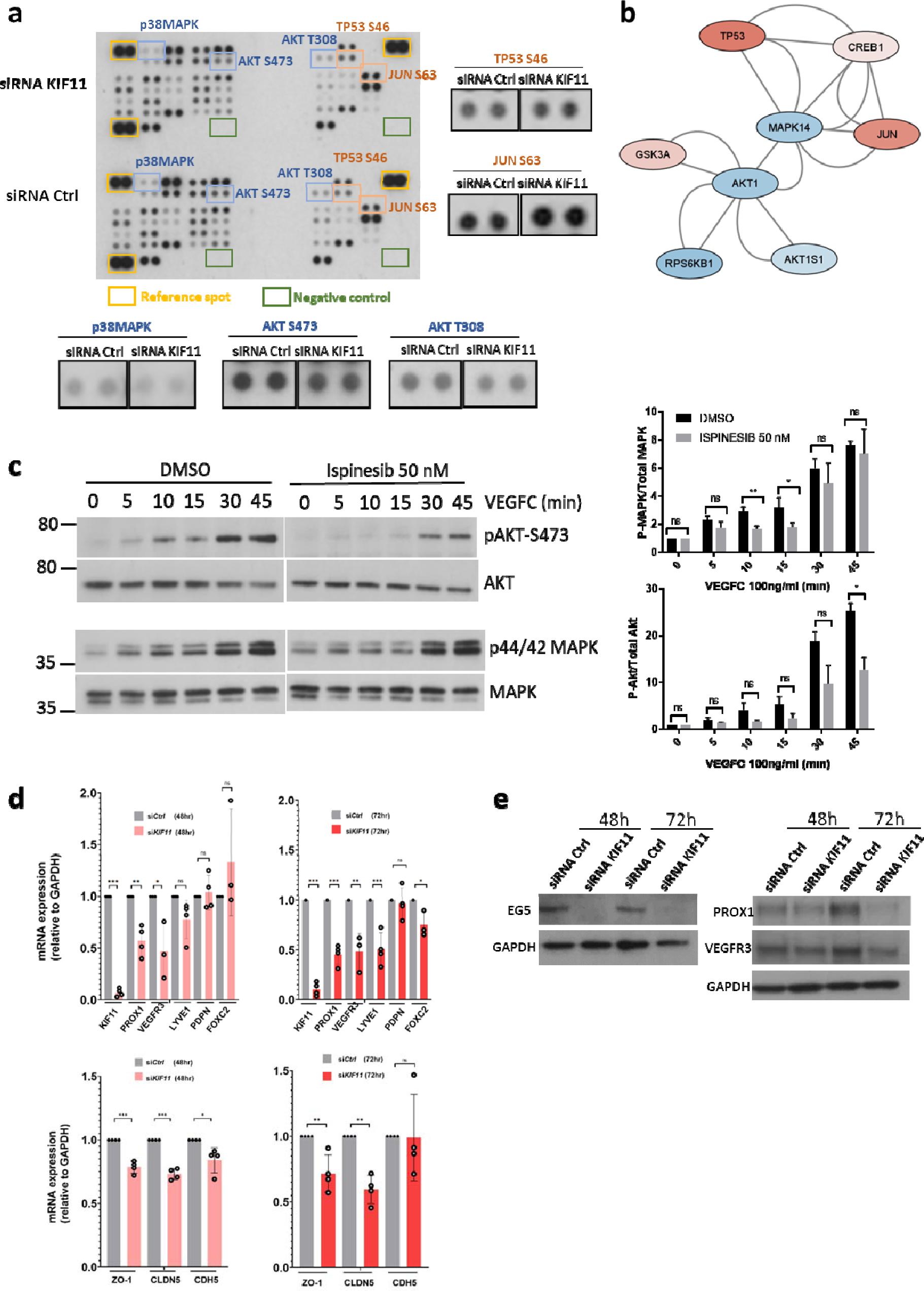
KIF11 deficiency alters MAPK and AKT signalling pathways and reduces expression of lymphatic cell identity and cell-cell junction markers. **(a)** Human Phospho-Kinase Array showing changes in phosphorylation levels of 43 kinases. Dots have been magnified for selected kinases with decreased (in blue) or increased (in red) phosphorylation. **(b)** Network pathway analysis using STRING and Cytoscape reveals known interactions and the directionallity of the phosphorylation change (blue, decreased and red, increased phosphorylation in siRNA KIF11 treated cells). **(c)** Effect of Ispinesib treatment (50nM) on AKT and MAPK phosphorylation in LECs over time after stimulation with VEGFC (100ng/mL). DMSO (vehicle) treated cells as control. Phosphorylation was analysed with anti-pAKT S473 and anti-p44/42 MAPK Thr202/Tyr204 (upper panels) and levels of AKT and MAPK protein expression with anti-AKT and MAPK antibodies (lower panels). Molecular mass markers (in kDa) is indicated on the left. Blots are quantified (on the right). (d) *VEGFR3*, *PROX1*, *Podoplanin (PDPN), FOXC2*, *LYVE1, ZO1, CLDN5 and CDH5* gene expression related to *GAPDH* in LECs treated with siRNA KIF11 for 48 and 72 hours analysed by qPCR. **(e)** Western blot analysis and quantification of PROX1 and VEGFR3 expression in LECs treated with siRNA KIF11 for 48 and 72 hours. GAPDH was used as control. (**c-e**) Bars represent mean relative expression ± SEM. (**c** and **e**) One representative image is shown from n=3-4 experiments. ns=not significant, *P<0.05, **P<0.01, ***P<0.001. Two-tailed unpaired Student’s t-test.

We focused our attention on the AKT and MAPK signalling pathways, that appeared downregulated after siRNA *KIF11* treatment, and are two of the major downstream effectors of tyrosine kinase receptor signalling including VEGFR3. LECs were pretreated overnight with DMSO (vehicle) or 50nM Ispinesib, and were stimulated the next day with 100ng/ml VEGFC for up to 45 minutes. VEGFC treatment activated the phosphorylation levels of MAPK and AKT both in DMSO and Ispinesib treated cells. Ispinesib treatment resulted in a significant reduction of MAPK phosphorylation at 10-15 min and AKT phosphorylation at 45 minutes post-VEGFC stimulation (**Fig. 7c**).

### KIF11 knockdown downregulated PROX1, VEGFR3 and cell junction molecule expression

PROX1 is a master regulator of LEC specification^37^ regulating genes important for maintenance of LEC identity such as VEGFR3 and Podoplanin^38,39^ and transcription factors essential for lymphatic valve morphogenesis like FOXC2^40^. To determine whether downregulation of lymphatic markers might also contribute to the lymphatic defects observed with *KIF11* haploinsufficiency, we transfected siRNA *KIF11* for 48 and 72 hours in LECs and investigated the effect on the expression of well-characterised lymphatic endothelial markers. After 48 hours we detected approximately 50% downregulation of *VEGFR3* gene expression and *PROX1* both at mRNA (**Fig. 7d**) and protein (**Fig. 7e**) levels. Other lymphatic identity markers such as *FOXC2* and *LYVE1* showed significant reduction after 72 hours whereas the gene expression of Podoplanin remained unaffected (**Fig. 7d**).

Lymphoscintigraphy (**Fig. 1a-c**) revealed a lack of interstitial fluid uptake in MLC patients suggesting the initial lymphatic capillaries may be dysfunctional. The permeability of the lymphatic capillaries is critical for functional fluid uptake and is controlled by specialised junctions^41^ and the importance of this specialized junctional organization has already been probed in several animal models^42–44^. Since it is crucial to understand the role of EG5 in controlling lymphatic junction architecture and permeability, and its contribution to lymphatic (dys)function (e.g. failure of fluid uptake in lymphatic capillaries), we looked next at the impact of *KIF11* depletion on the expression of cell junction molecules and found downregulation of ZO-1 and Claudin-5 but not VE-Cadherin (**Fig. 7d**).

## Discussion

Autosomal dominant pathogenic variants in *KIF11* (EG5) are causative of microcephaly-lymphedema-chorioretinopathy (MLC) syndrome, a condition that presents with a variable spectrum of central nervous system, lymphatic and ocular developmental anomalies^1,14^. Since the association of *KIF11* with MLC^3^, we have aimed to understand how EG5 mutations account for the lymphatic phenotype presented in this pleiotropic disease. Very little was previously understood about the underlying mechanisms leading to lymphatic failure, partly due to the limited knowledge about the role of *KIF11* in the development and maintenance of the lymphatic system. A better understanding of the signalling pathways and molecules that regulate lymphatic development and physiology in the context of *KIF11* pathogenesis is essential to enable the development of more targeted and accurate treatments for MLC patients.

In this study we were examining the disease mechanism(s) causing lymphedema in patients with mutations in *KIF11*, as well as investigating the molecular and cellular interactions by which *KIF11* may regulate lymphangiogenesis. We have attempted this through; 1) investigations on the clinical characteristics and lymphatic function of patients with MLC, by lymphocintigraphy and immunohistochemystry of intestinal biopsy; 2) the use of patient-derived cells to study the expression levels of *KIF11*; 3) the analysis of *KIF11* expression during embryonic lymphatic development both in mouse and human; 4) the generation of a MLC zebrafish model; and 5) an examination of the impact of *KIF11* happloinsufficency on VEGFR3 signalling and lymphatic identity using primary lymphatic endothelial cells as an *in vitro* model.

As previously reported, lymphedema in MLC patients is mostly bilateral, affecting the lower limbs. To determine whether the fault in the lymphatic function is at the initial lymphatic capillaries or at the collector level we performed lymphocintigraphy in three MLC patients carrying two stop-gain pathogenic variants in *KIF11*. Images showed a typical functional aplasia mechanism where the tracer (or most of it) was retained at the injection points, meaning a failure or reduction of the ability of the initial lymphatics. This pattern is similar to that observed in Milroy patients with pathogenic variants in *VEGFR3*^45^ and suggests that the fault is in the initial lymphatic capillaries.

One patient presented with intestinal lymphangiectasia, and the staining of an intestinal biopsy revealed reduced Podoplanin positive lymphatic vessels. Reduced number of dermal lymphatic vessels has also been observed in skin biopsies from Milroy patients^46^. Some mechanisms suggested to cause lymphangiectasia and impaired lacteal function include defective LEC polarity, valve maldevelopment, lymphatic vasculature hyperplasia and junctional destabilization^47,48^. Due to the critical role of the lacteals in fat absortion and digestion^49^ the lymphangiectasia observed is likely to explain the intestinal problems in this patient. Further analysis of several blood and lymphatic markers together with junctional proteins could reveal additionally structural and/or functional alterations. Only one case of intestinal lymphangiectasia had been observed in MLC before^14^, therefore, our finding confirms that intestinal lymphangiectasia should be considered a feature of MLC, albeit not typical, and clinicians should perform investigations accordingly. This finding also provides new suggestions for the clinicians for patient care (e.g. prescription of a low-fat diet for intestinal lymphangiectasia in MLC patients).

Scientists have searched for the molecular disease mechanisms of MLC, suggesting mutations would most likely result in loss of function of *KIF11* due to nonsense-mediated mRNA decay, premature truncation of the protein or splicing abnormalities^14^. In this study we looked at the impact of *KIF11* mutations on mRNA and protein expression levels in lymphoblastoid transformed cells isolated from the blood of patients (together with blood and saliva samples) confirming that the two stop-codon variants investigated resulted in a reduction but not total loss of the mutated mRNA. As expected, qPCR and western blot analysis revealed an approximately 50% reduction of *KIF11* expression and in one case a shorter truncated EG5 protein was detected. A synonymous variant in *KIF11* investigated in this study caused cryptic splicing which led to exon skipping. Others showed a similar effect for the synonymous variant c.2922G>T^50^. Therefore, we conclude that *KIF11* haploinsufficiency is the likely underlying key disease mechanism in MLC.

We believe that to be able to explain the pathophysiology of MLC it is neccesary to understand the association of *KIF11* with the development of the lymphatic system, particuarly since the expression pattern of *KIF11* in the lymphatics during development and adulhood has been poorly investigated. The early onset of lymphedema in MLC also suggests that the interaction has occurred early in development. We found *KIF11* expression associated with the development of lymphatic structures both in human and mouse embryos using RNA scope *in situ* hybridyzation. Co-expression of *KIF11* was detected together with lymphatic endothelial markers such as PROX1, VEGFR3 or Podoplanin. Interestingly we found association of *KIF11* regulatory regions with the lymphatic transcription factors PROX1, FOXC2 and NFATC1. Another intriguing finding was to discover *KIF11* mRNA signal higher in dermal LECs compared to that of the pTD/lymph sacs, which could indicate a distinct role for *KIF11* (EG5) in discrete lymphatic vessel beds. We could speculate that the relative prominence of *KIF11* expression in dermal LECs could relate to the functional aplasia of the initial dermal lymphatics observed in lymph scans from MLC patients.

Heterozygous mutant mice generated with a genetrap insertion in Eg5 appear phenotypically normal. In contrast, embryos homozygous for the Eg5 null allele displayed signs of a proliferation defect suggesting Eg5 is essential for cell division during early development^8^. In another study, Chauviere and colleagues demonstrated that heterozygous mice are healthy, fertile, and show no detectable phenotype, whereas Eg5(*/*) embryos die during early embryogenesis^7^. None of these studies attempted to look at the impact of *KIF11* deficiency in the lymphatic system.

The generation of *in vivo* models to further investigate *KIF11*’s role in lymphatic function has proved a difficult journey, most probably because of the critical role of EG5 in mitosis and the need for depleting Eg5 early during embryo development to study its role in lymphatic development. Analysis of scRNA-seq data of wildtype zebrafish cell populations at different developmental stages confirmed the expression of kif11 associated with highly proliferative lymphatic and blood endothelial precursors during zebrafish embryonic development. This could justify why we observed severe defects in cell survival and development that lead to early embryonic lethality. Therefore, even with the introduction of a deletion mimicking the variants observed in some MLC patients, we do not see a comparable phenotype in zebrafish. It is possible that *kif11* function may be compensated by another kinesin motor protein or that divergence in kinesin motor protein functions is at play in zebrafish. Nevertheless, this work demonstrates an essential role in early development for *kif11* and its requirement for embryo survival in zebrafish.

We then turned to *in vitro* models to investigate the possible role of *KIF11* in lymphatic endothelial cells. We focussed on cell migration since the co-expression of *KIF11* and *VEGFR3* mRNA in the primordial lymphatic developing sacs could point to a possible participation of EG5 in the proliferative and migratory activity of the LEC precursor cells leaving the cardinal vein. Moreover, previous publications have highlighted this non-mitotic role linking KIF11 with migration in several cell types^10,51,52^. Our experiments suggest that LEC migration is affected by EG5 inhibition, which could be explained by the importance of microtubules in cell locomotion. Studies show how suppression of microtubule dynamics restrained forward progression and impaired directionality^53^ and is dependent on the right level of kinesin expression^54^.

VEGFR3 is the master regulator of lymphatic function and development and we have previously shown that Milroy patients present with reduced size of lymphatic capillaries in their skin^46^. We have also observed reduced Podoplanin positive structures in the intestinal biopsy of an MLC patient. The remarkable similarities between the bilateral, congenital pedal lymphedema seen in Milroy disease (caused by *VEGFR3* mutations) and MLC (caused by *KIF11* mutations) point to a plausible convergence of both molecules in the same functional signalling axis. The co-expression of *KIF11* and *VEGFR3* positive vessels during human embryonic development supports this hypothesis of a common functional pathway.

We have observed a downregulation of AKT and MAPK phosphorylation after EG5 inhibition. Further research will be needed to investigate the specificity of VEGFC on the observed effect on AKT and MAPK phosphorylation after EG5 inhibition. A decrease of VEGFR3 signalling through loss of AKT activation could have a detrimental effect on the migratory behavour of LECs, as it has been shown that hyperactivation of PIK3CA confers a migratory LEC phenotype through increasing P-AKTSer473^55^. This finding opens a new avenue for future studies to investigate the role of EG5 in the PIK3/AKT pathway and the possibility of repurposing AKT activators (e.g. SC-79) as a potential treatment of MLC^56^.

*KIF11* knockdown downregulated PROX1, VEGFR3 and cell junction molecule expression. Other genes associated with primary lymphatic anomalies have been shown to regulate cellular junctions and permeability. For instance, EphrinB2/EPHB4 defective mice show impaired junctions^42^ which could explain the edema observed. Interestingly, other kinesins have been shown to mediate VEGFR2 cell membrane recycling through Rab11, regulating vascular permeability^57^. Analysis of skin biopsies from MLC patients could confirm junctional loss of integrity as the disease mechanism and possibly shed light on the functional aplasia observed.

In conclusion, we confirm *KIF11* haploinsufficiency in MLC patients as the disease mechanism and that EG5 could already play a role during embryonic lymphangiogenesis as its inhibition impairs lymphatic endothelial cell functions. We speculate whether the lymphatic impairment resulting in functional aplasia (and intestinal lymphangiectasia) observed in MLC patients could be a consequence of dysregulated VEGFR3 signalling. This would impact on the control of cell communication and signalling cascades orchestrating cell proliferation and migration during lymphangiogenesis. Our data provide the first insights into the study of *KIF11*(EG5) in lymphatic function, and future studies should elucidate the specific disease mechanisms connecting *KIF11* pathogenic variants and lymphedema and the possibility of exploring AKT activators as potential treatment for MLC patients.

## Acknowledgements

We extend our thanks to the patients and their families, and to the ‘KIF11Kids Association’ in Germany. This work was supported by the British Heart Foundation (BHF) [SP/13/5/30288 and FS/15/39/31526] and a joint grant from the Medical Research Council (MRC) and the British Heart Foundation (BHF) [MR/P011543/1 and RG/17/7/33217]. SMA was supported by a research fellowship from the Rosetrees Trust [StGeorges-21\2] and St George’s Hospital Charity [RES 20-21 003]. We thank Dr Femke Piersma (RMK Winnenden) and Dr Tränkenschuh (Robert Bosch Klinikum Stuttgart) for providing the patient intestinal biopsy, H&E and immonohistochemistry images and interpretation, and Dr Sue Heenan for the lymphoscintigraphy images. The human embryonic and fetal material was provided by the Joint MRC / Wellcome (MR/R006237/1) Human Developmental Biology Resource (www.hdbr.org). We thank Dr Anna Oszmiana (University of South Australia) for sharing the ChIP-seq data. We also thanks Dr Michelle Meier (University of Melbourne) and Dr Kazuhide S. Okuda (University of Melbourne) for generating figures and providing insights for the zebrafish work, respectively.

For the purpose of open access, the author has applied a Creative Commons Attribution (CC BY) licence to any Author Accepted Manuscript version arising.

## Author contributions

Conceptualization: S.J., P.M., S.M., S.M.A., P.O.; Project administration: S.M.A., P.O.; Investigation: K.O., I.M.C., R.Y.B., R.B., S.U., N.H., E.S., A.A., C.K., K.G., T.M., S.M.A.;, Formal analysis: KO, S.D., D.G., B.H., T.M., R.H., S.M.A., P.O.; Resources: M.O., D.W., A.E., K.K., K.G., B.H., R.H., S.M., P.O.; Writing – original draft: K.O., S.M.A., P.O.; Writing – review & editing: All authors. Supervision: S.J., P.M., B.H., T.M., R.H., S.M., S.M.A., P.O.; Funding acquisition: S.J., P.M., S.M., P.O.

## Data availability

Data are available in the article itself and its supplementary materials. Source Data is provided in pdf format accompanying this manuscript available online. Data used to generate Fig 3b and Fig 5 a-d were derived from sources in the public domain. Raw data is available from the corresponding authors upon reasonable request.

