## Supplementary information for "Insights into *KIF11* pathogenesis in Microcephaly-Lymphedema-Chorioretinopathy syndrome: a lymphatic perspective"

**Supplemental Tables and Figures and Video legends**

**Table S1: Clinical summary of MLC patients with confirmed *KIF11* variants.**

**Table S2: Annotation of the four *KIF11* variants identified in this study.**

**Table S3: Oligonucleotides (5’- 3’) used for RT-qPCR in Fig 7d.**

**Fig. S1: Pedigrees of MLC families. (a)** The pedigrees of six families featuring *KIF11* variants are shown. The probands are marked with arrows. The stars indicate the individuals who had a lymphoscintigraphy scan. The question marks represent individuals with unknown disease or condition status. The cross indicates the individual who underwent biopsy.

**Fig. S2: *KIF11* loss-of-function in *in vitro* models in human dermal lymphatic endothelial cells. (a)** EG5 protein levels were assessed by western blot 24 hours post-transfection with siRNA KIF11 or siRNA Ctrl. GAPDH used as loading control. One representative image is shown from n=3 experiments. Data represent mean ± SEM. ****P<0.0001. Two-tailed unpaired Student’s t-test. **(b)** 24 hours after transfection with siRNA KIF11 or siRNA Ctrl, proliferating LECs were labelled with Alexa Fluor 647 after EdU incorporation and Hoechst 33342 for the staining of the total nuclei. Data represents mean ± SEM (n=3 biological replicates with 3 technical replicates each). **P<0.01. two-tailed unpaired student’s t-test. **(c)** In DMSO (vehicle) treated cells, EG5 (green) and α-tubulin (red) co-localize in the bipolar spindle poles. Co-localization is lost in LECs treated with Ispinesib 50nM and 5nM (but not 0.5nM) and show abnormal monopolar spindle poles (arrow heads). Scale bar 10 µm, n=1. **(d)** CellTracker™ Red CMTPX staining of live 3D spheroids. At the end of the 3D spheroid assay (24 hours) where spheroids exposed to DMSO control or various concentrations of Ispinesib (50,100,200nM), spheroids were stained for 2 hours with non-toxic ThermoFisher CellTracker. Spheroid images were captured by EVOS M5000 and fluorescence intensity of dye was measured (using ImageJ). Fluorescence dye directly indicates viable cells as the dye can only be retained in living cells which then can transfer to daughter cells but not adjacent cells. Representative images are shown from n=2 biological experiments from total of 10 spheroids.

**Video S1: A label-free high content imaging system to capture the *in vitro* wound healing process of Lymphatic Endothelial Cells (LECs), in (a) DMSO control and (b) 50nM Ispinesib treated cells**. Phase contrast video (black=background; white=LECs) of *in vitro* wound healing process of human primary lymphatic endothelial cells (LECs) being exposed to DMSO control or 50nM ispinesib. MP4 video was created by capturing wound healing process every 10 minute intervals for 15 hours. (25.00 frames/sec; frame width=768, height=768).

**Table S1. Clinical summary of MLC patients with confirmed *KIF11* variants.**

| **Family** | **ID** | **Sex** | **Current Age range (years)** | **Nucleotide variant** | **Bilateral lower limb lymphedema** | **Age of onset** | **Systemic involvement** | **Head circumference (SD)** | **Retinal changes or other eye features** | **Dysmorphic** | **Learning difficulties** |
| --- | --- | --- | --- | --- | --- | --- | --- | --- | --- | --- | --- |
| F1 | II.1 | F | 20-29 | c.1159C>T | Y | Birth | N | -8.0 | Y | Y | Mild |
| F1 | I.2 | M | 50-59 | c.1159C>T | Minimal |  | N | -2.0 | N | N | Mild |
| F2 | II.1 | F | 10-19 | c.1039_1040del | Y | Birth | N | -5.7 | Y | Y | Mild |
| F2 | I.2 | M | 40-49 | c.1039_1040del | N |  | N | -2.0 | N | Mild | Mild |
| F3 | II.1 | M | 30-39 | c.2680C>T | Y | Birth | N | -3.2 | (Y) | N | Moderate |
| F3 | II.2 | M | 30-39 | c.2680C>T | Y | Birth | N | -3.2 | (Y) | N | Autism |
| F4 | II.1 | M | 0-9 | c.2922G>A | Y | Birth | Y | -4.5 | Y | Y | Mild |
| F5 | II.1 | M | 20-29 | c.2922G>A | Y | (Birth) |  | na | Y |  | Mild |
| F6 | II.1 | F | 10-19 | c.2922G>A | Y | <3 mth | N | -4.8 | Y | Y | N |

F, female; M, male; mth, month; N, no; na, microcephaly confirmed but details not available; SD, standard deviation; Y, yes; ( ), unconfirmed clinical finding.

**Table S2: Annotation of the four *KIF11* variants identified in this study.**

| **Family ID** | **Genomic coordinates (GRCh38)** | **Nucleotide change** | **Predicted protein change** | **CADD Phred score** | **Mutation Taster prediction** | **ACMG classification** | **Reported by** |
| --- | --- | --- | --- | --- | --- | --- | --- |
| F1 | chr10:92621415 | c.1159C>T | p.R387* | 35 | Disease causing | Pathogenic (PVS1, PM2, PP1) | Ostergaard et al., 2012 |
| F2 | chr10:92616741-92616742 | c.1039_1040del | p.L347Efs*8 | 33 | Disease causing | Pathogenic (PVS1, PM2, PP1) | Ostergaard et al., 2012 |
| F3 | chr10:92648344 | c.2680C>T | p.Q894* | 37 | Disease causing | Pathogenic (PVS1, PM2, PP1) | This study |
| F4 | chr10:92649986 | c.2922G>A | p.939_974del | 22.8 | Disease causing | Likely pathogenic (PP3, PM2, PS2) | This study |
| F5 |  |  |  |  |  |  |  |
| F6 |  |  |  |  |  |  |  |

Genomic coordinates (GRCh38), nucleotide and predicted protein changes are summarised. The variants were not present in gnomAD v2.1 or v3 databases and they are predicted to be pathogenic by two prediction tools (CADD and MutationTaster). ACMG classify the protein truncating variants as pathogenic or likely pathogenic.

**Table S3: Oligonucleotides (5’- 3’) used for RT-qPCR in Fig 7**d.

| KIF11 | Forward | AGCAAGCTGCTTAACACAGTT |
| --- | --- | --- |
|  | Reverse | CCTTCTTACGATCCAGTTTGGAA |
| GAPDH | Forward | CAAGGTCATCCATGACAACTTTG |
|  | Reverse | GGGCCATCCACAGTCTTCTG |
| VEGFR3 | Forward | TGCACGAGGTACATGCCAAC |
|  | Reverse | GCTGCTCAAAGTCTCTCACGAA |
| PROX1 | Forward | TACGCACGTCAAGCCATCAA |
|  | Reverse | CAGGAATCTCTCTGGAACCTCA |
| ZO-1 | Forward | CGGTCCTCTGAGCCTGTAAG |
|  | Reverse | GGATCTACATGCGACGACAA |
| CLDN5 | Forward | CTCTGCTGGTTCGCCAACAT |
|  | Reverse | CACAGACGGGTCGTAAAACTC |
| CDH5 | Forward | GATCAAGTCAAGCGTGAGTCG |
|  | Reverse | AGCCTCTCAATGGCGAACAC |
| PDPN | Forward | GGGAAGGTACTCGCCCTAAAG |
|  | Reverse | CACGGGTCATCTTCTCCCAC |
| FOXC2 | Forward | GGGGACCTGAACCACCTC |
|  | Reverse | AACATCTCCCGCAGTTG |
| LYVE1 | Forward | GCTTCAGCCTGGTGTTGCTT |
|  | Reverse | GCCGGCCAAACTTAGTCCCA |

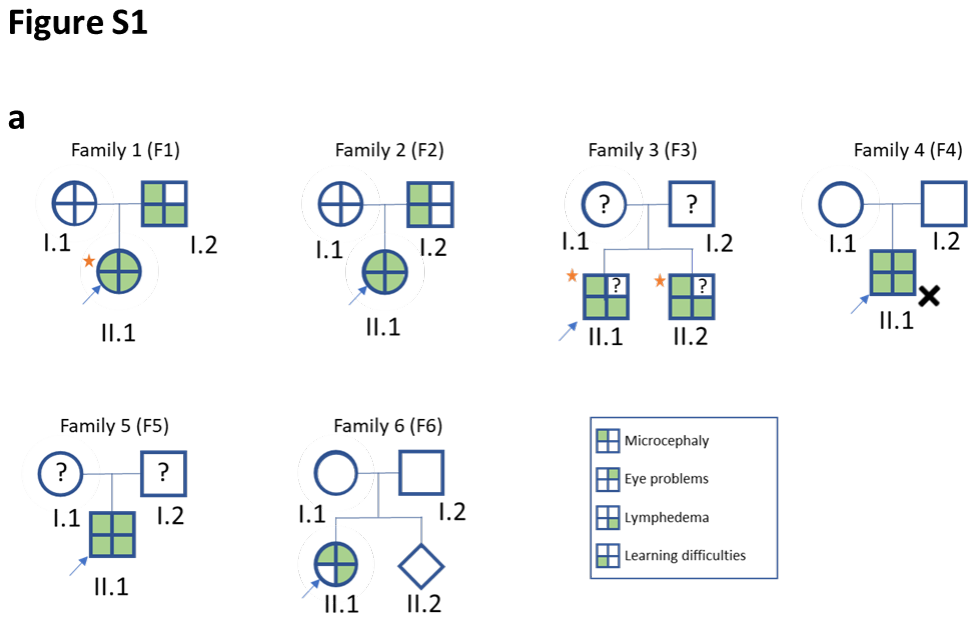

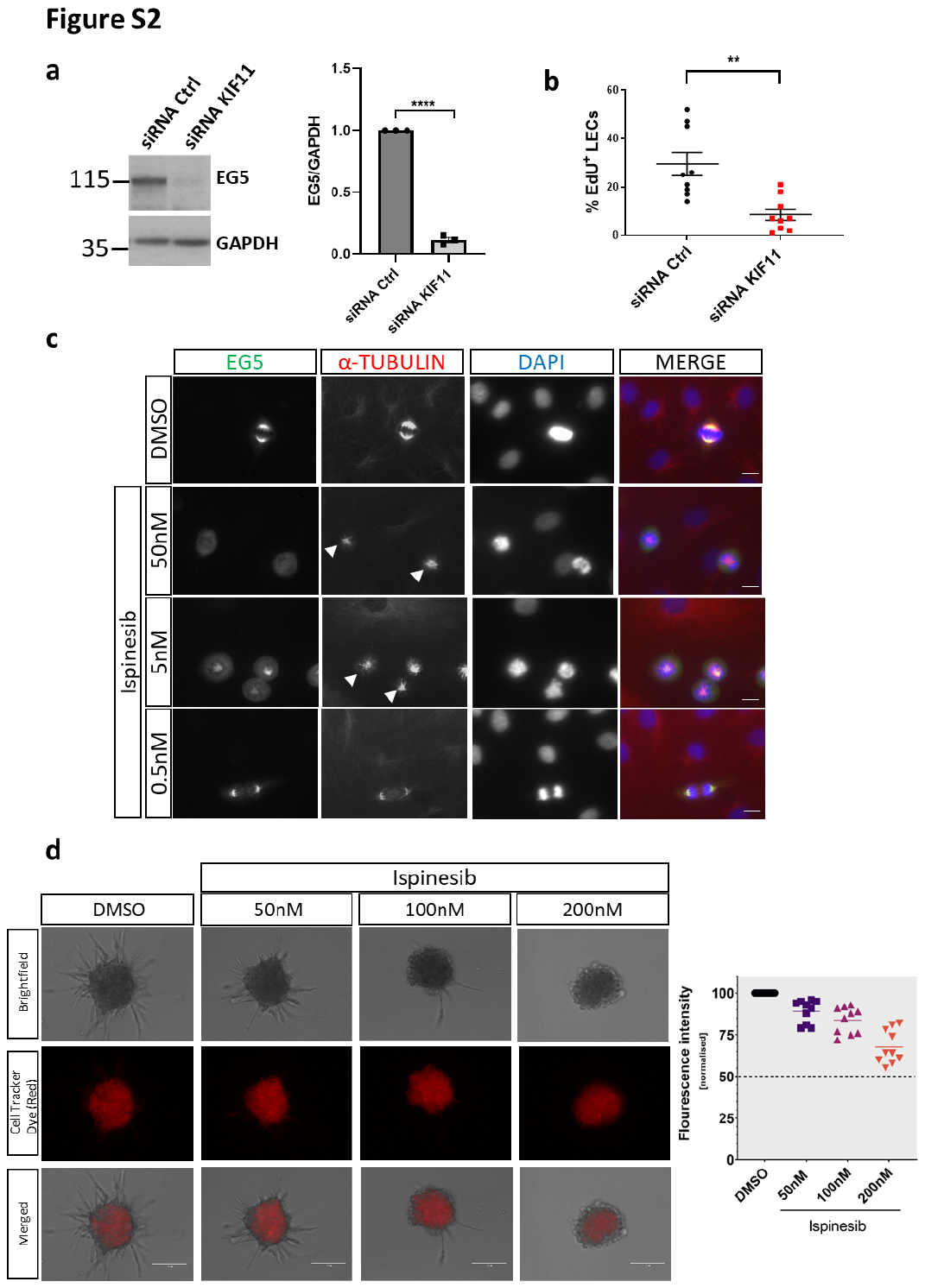

**Supplementary Materials and Methods**

*Lymphoscintigraphy*

Three patients underwent lower limb lymphoscintigraphy, which is the imaging of the lymphatic system. This was performed according to standard local procedure by injecting a radioactive isotope (technetium-99m-nanocoll) into the web spaces between the toes^1^ and imaging the uptake into the inguinal lymph nodes after 2h with a gamma camera. Quantification figures 2h post-injection were calculated as percentage (%) of tracer retention within the feet, and tracer uptake in the ilioinguinal nodes.

*Mouse embryo collection*, *histology and immunofluorescence*

Wild-type mouse embryos of the C57BL/6 genetic background were analyzed at developmental stages, E10.5 to E13.5. Day of embryonic development (E) was determined by vaginal plug appearance (E0.5). Laboratory mouse work was approved by German Federal Authorities (LaGeSo Berlin) under the license number ZH120. All animal procedures were in accordance with institutional, state, and government regulations. Embryos collected at different developmental stages between E10.5 and E13.5 were fixed in 4% PFA/PBS at room temperature for 2-4 hours, depending on sample size. After fixation, samples were washed twice in PBS before incubation in an increasing sucrose series (5%, 10% and finally 15% (w/v) sucrose/PBS), each for an hour or until the embryos sank to the bottom of the tube. Finally, embryos were incubated in a 1:1 solution of 15% (w/v) sucrose/PBS and Tissue-Tek® O.C.T. Compound (94-4583, Sakura) for 1 hour at room temperature. After transfer into Tissue-Tek® O.C.T. Compound fully-embedded embryos were snap-frozen in a chilled ethanol bath and stored at -80°C until sectioning. For standard histology, 5 μm cryo-sections of embedded embryos were fixed in 4% PFA/PBS for 15 min, washed in PBS, and stained with Hematoxylin and Eosin for 1 minute. After mounting with Entellan (Sigma-Aldrich, 1.07960), images were captured using a Zeiss Axio Observer Z7 (20× air, NA = 0.8) epi-fluorescence microscope.  For standard immunofluorescence, 10 μm cryo-sections of embedded embryos were fixed in ice-cold methanol for 15 min, washed in PBS and blocked (10% chicken serum, 0.3% Triton X-100 in PBS) for 1 hour min at room temperature. After blocking, tissue sections were incubated for 1 hour with primary antibodies (diluted in 1% BSA, 1% chicken serum, 0.3% Triton X-100 in PBS), washed thrice in PBS-T (0.1% Tween20 in PBS) and incubated in Alexa dye–conjugated secondary antibodies. Finally, after 3 washing steps in PBS-T, samples were stained with Hoechst to visualize cell nuclei. After mounting in Fluorescence Mounting Medium (S3023, Dako Agilent, Santa Clara, US), samples were imaged using a Zeiss LSM 980 confocal microscope (25x oil, NA = 0.8). The following antibodies were used: Hoechst 33342 solution (62249, Waltham, MA, USA), rat monoclonal IgG2a anti-mouse PECAM-1 (102502, Biolegend, San Diego, CA, USA), rabbit polyclonal IgG anti-mouse PROX-1 (102-PA32AG, ReliaTech, Wolfenbüttel, DE), goat polyclonal IgG anti-mouse VEGFR3 (AF743, R&D systems, Minneapolis, MN, USA), donkey polyclonal anti-mouse IgG Alexa Fluor 488 (A21208, Invitrogen, Waltham, MA, USA), donkey polyclonal anti-rabbit IgG Alexa Fluor 568 (A10042, Invitrogen, Waltham, MA, USA), donkey polyclonal anti-goat IgG Alexa Fluor 647 (A32849, Invitrogen, Waltham, MA, US).

*RNAscope in situ hybridization in mouse embryos*

Mouse embryos were collected at different developmental stages between E10.5 and E14.5. They were fixed for 4 hours in 4% PFA/PBS at room temperature. The embryos were washed twice in PBS before incubation in an increasing sucrose series (5%, 10% and finally 15% (w/v) sucrose (Roth/PBS) each for an hour or until the embryos sank to the bottom of the tube. Finally, the embryos were incubated in 15% (w/v) sucrose/PBS and Tissue-Tek® O.C.T. Compound (94-4583, Sakura) in a 1:1 solution before embedding the embryos in Tissue-Tek® O.C.T. Compound in a chilled ethanol bath and stored at -80°C for sectioning. The embryos were cut into 5 μm thick sections on slides for RNAscope. Simultaneous RNA *in situ* hybridization was performed using the RNAscope® technology (Advanced Cell Diagnostics [ACD]) and the following probes specific for Mm-VEGFR3 (Cat. No. 481371, ACD), Mm-ERG (Cat. No. 546491-C2, ACD) and Mm-KIF11 (Cat. No. 819941-C3, ACD) on five μm sections of the mouse embryos. RNAscope probes were purchased and designed by ACD. The RNAscope® assay was run on a HybEZ™II Hybridization System (Cat. No. 321720, ACD) using the RNAscope® Multiplex Fluorescent Reagent Kit v2 (Cat. No. 323100, ACD) and the manufacturer’s protocol for fixed-frozen tissue samples with target retrieval on a hotplate for 5 minutes. Fluorescent labelling of the RNAscope® probes was achieved by using OPAL 520, OPAL 570 and OPAL 690 dyes (Cat. No. FP1487001KT, Cat. No. FP1488001KT, Cat. No. FP14970014KT, Akoya Biosciences, Marlborough, MA, USA) and stained sections were scanned using an LSM 980 with Airyscan 2 at various magnifications (Carl Zeiss AG, Oberkochen, DE). For quantification of mRNA signal RNAscope probes specific for VEGFR3 and KIF11 were used. The number of signal-positive spots per area (25µm^2^) for each probe was determined by manual counting. For each image up to 8 areas were analysed using ImageJ.

*ChIP-Seq data analysis*

FOXC2, NFATC1 and PROX1 ChIP-Seq data was downloaded from NCBI sequence read archive (SRA) under accession number SRP191993, and GATA2 from the European Nucleotide Archive under accession number [PRJEB9436](http://www.ebi.ac.uk/ena/data/view/PRJEB9436). Data was processed using trim galore (<https://www.bioinformatics.babraham.ac.uk/projects/trim_galore/>), fastqc (<http://www.bioinformatics.babraham.ac.uk/projects/fastqc>) and aligned to human reference genome (b38) with Bowtie2 (v2.5.0)^2^. Alignments were visualized using the Integrative Genomics Viewer (2.16)^3^.

*RNA isolation from LECs and RT-qPCR*

RNA was isolated from LECs with the miniRNA extraction kit (Qiagen). 500 ng of total RNA was used as template for cDNA generation using Superscript III and Oligo dT (Invitrogen). Oligonucleotides are listed in **Table S3**, and all efficiencies were confirmed above 95% in a standard curve analysis. Real-time PCR was performed using SYBR Green (Promega) and Bio-Rad CFX96 thermocycler. The results were analysed using Bio-Rad CFX Manager Software version 2.0. Quantification was done with the ΔΔCT method to calculate fold-differences with the reference gene *GAPDH*.

*Immunofluorescence and Western blot*

For immunofluorescence LECs grown on fibronectin-coated glass coverslips were fixed in 4% PFA for 15 minutes and permeabilized with 0.5% Triton X-100 for 5 minutes at room temperature. Blocking was done with 1% BSA for 1 hour before incubation with primary antibodies (EG5 1:50 ab72413, Abcam and α-tubulin (DM1A) 1:200 T9026, Sigma) overnight at 4ºC. Coverslips were then incubated with secondary antibodies (anti-rabbit Alexa Fluor 488 1:500 A-21206 and anti-mouse Alexa Fluor 555 1:500 A31570, Life Technologies) for 1 hour at room temperature, DAPI (Sigma) for 5 minutes and mounted in Vectashield (Vector Laboratories). Images were taken with a Zeiss Axiovert 200M in the St George’s University of London (SGUL) Image Research Facility.

For western blot LECs or lymphoblastoid cell lines were harvested in lysis buffer (20 mM Tris pH 7.5, 150 mM NaCl, 0.5%Triton X-100) supplemented with protease and phosphatase inhibitors (Sigma-Aldrich). After clarification by centrifugation, protein lysates were separated by SDS-PAGE and transferred to PVDF-P membranes that were blocked with 5% milk or BSA in PBS. Immunoblot analysis was performed with the following primary antibodies: anti-human EG5 (1:1000, ab72413-Abcam, CC10014-Cell Applications, NB500-181-Novus Biologicals), anti-human GAPDH (1:10000, MAB374, Merck Millipore), anti-human PROX1 (1:1000, AF2727, Biotechne), anti-human VEGFR3 (1:50, MAB3757, Merck Millipore), anti phospho-Akt (Ser473) (1:1000, #4051, Cell Signalling), anti Akt (1:1000, #9272, Cell Signalling), anti-phospho-p44/42 MARPK (Erk1/2) (Thr202/Tyr204) (1:1000, #9106, Cell Signalling), anti p44/42 MARPK (Erk1/2) (1:1000, #9102, Cell Signalling). Secondary antibodies were HRP-conjugated and chemiluminescence detection was performed with ECL™ (GE Healthcare). Protein expression was quantified by densitometry and normalized against GAPDH using Image J.

*Transwell migration assay*

LECs were serum starved for 16 hours prior to seeding into 8 μm cell culture transwell inserts (BD biosciences) placed in 24 well plates (Santacruz, USA) and coated with fibronectin. Prior to seeding cells, inserts were filled with PBS (with Ca^2+^ and Mg^2+^) and incubated for an hour. 5x10^4^ cells per insert were plated in endothelial medium MV2 and the same medium supplemented with 100ng/mL VEGFC was added to the lower chamber. After 10 hours, inserted membranes were fixed in methanol at -20^º^C and nuclei stained with DAPI. After removing residual cells in the upper chamber with a cotton swab, nuclei on the lower surface of the membrane were counted using an EVOS M5000 digital inverted microscope (10X). An average count of 5 different fields of view were quantified.

*Wound healing assay*

LECs were seeded within two-well culture inserts (ibidi) attached to fibronectin-coated glass bottomed plates at a density of 34,000 cells/insert chamber. After 2 hours, the insert was removed, and cells were washed twice with PBS containing Ca^2+^ and Mg^2+^ before exposure to either vehicle (DMSO 0.01%) or 50nM Ispinesib in endothelial cell growth medium MV2 with 100ng/mL VEGFC.

*Single cell migration tracking*

Quantitative phase image microscopy assays were performed using a Livecyte microscope (Phase Focus Limited, UK) according to the manufacturer’s instructions. In brief, LECs were plated and treated as described for wound healing assay (two independent experiments [#1 and #2] were performed using two distinct LEC single-donor batches). Images of wound closure were acquired every 10 minutes for 15 hours using a 10x objective lens at 37°C and 5% CO_2_. Analysis of tracks to evaluate cell motility and speed was implemented using CellTrackR (v1.1.0)^4^ . Analysis of tracks was performed over time frames (50 ≥ t ≤ 100). Star plots were generated with the CellTrackR normalizeTracks feature selecting data with a minimum track length of 40. Thirty tracks were randomly selected to be displayed from the full set using the R runif function. To reduce potential bias^5^ velocity was calculated for each step for all cells and tracks. For experiment #1 the analysis was based on 2,270 control and 1,991 treatment tracks (44,686 and 43,613 steps). For experiment #2 the analysis was based on 6,676 control and 9,927 treatment tracks (89,804 and 97,713 steps). The difference between the distributions of speeds was assessed using the Wilcoxon-rank-sum test implemented in R. As an alternative metric for investigating cell motility, we also assessed the number of cells entering the scratch-window and the area of cells / scratch-window area across timeframes (0>t<200). The scratch window areas were defined by x = (300-500) for experiment #1 and x = (400-600) for experiment #2. One rogue cell with area >7,000µm² was removed from the analysis.

*Spheroid sprouting assay*

To generate cell spheroids (optimally ~750 cells per spheroid) LECs were trypsinized and suspended in growth factor supplemented MV2 medium without VEGFC and seeded in ultra-low adhesive round-bottomed 96-well plates (Cat 4515, Greiner, Frickenhausen, Germany). After 24 hours at 37^º^C (5% CO2), spheroids were harvested. Collagen stock solution was prepared prior to use by mixing 8 volumes (2mg/ml, 4^º^C) of rat tail collagen type-1 (Santa Cruz, USA) with 1 volume of 10X HBSS (Gibco BRL, Eggenstein, Germany), and 1 volume of 0.2 N NaOH. Neutralised collagen solution was mixed with harvested spheroids (10-15 spheroids/condition). 150ng/mL VEGFC and 50nM Ispinesib were also added into a neutralised collagen and spheroids mix, before transferring to prewarmed tissue culture plates. After 30 minutes at 37^º^C to allow gel polymerization, 0.2mL MV2 medium containing growth factors was pipetted on top of each gel. After 6-24 hours, *in vitro* sprouting was quantitated digitally (EVOSM5000) by measuring the number of the sprouts per spheroid and the length of each sprout (calculated and presented as average sprout length per spheroid).

*Proliferation assay*

LECs were transfected with siRNA Control or siRNA *KIF11* and after 24 hours proliferating cells were labelled with the Click-iT™ Edu Cell Proliferation Kit for Imaging, Alexa Fluor™ 647 dye (C10340) following the manufacturer’s instructions. Images were taken at 20x magnification using the Nikon A1R inverted confocal microscope (St George’s University of London (SGUL) Image Research Facility). For the quantification of LEC proliferation, one picture from the center of each sample was captured and the total number of nuclei, as well as the number of EdU positive cells, were counted using the Nikon NIS-Elements C software.

stylefix
